# Ethnic and sex inequalities in premature coronary artery disease across disaggregated South Asian and Black subgroups in England: a population-based cohort study

**DOI:** 10.64898/2026.08.03.26359414

**Authors:** Nalin Natarajan, Simon R Parker, Sam Quill, Sadaf Diamondali, Krishnaraj Rathod, Fizzah Choudry, Abhishek Joshi, Jorgen Engmann, A Floriaan Schmidt, Aroon D. Hingorani, Sophie Eastwood, Nishi Chaturvedi, Riyaz S. Patel

**Affiliations:** Institute of Cardiovascular Sciences, UCL, 222 Euston Road, London, United Kingdom; NIHR University College London Biomedical Research Centre, University College London and University College London Hospitals NHS Foundation Trust, London, United Kingdom; Bart’s Heart Centre, St Bartholomew’s Hospital, London, United Kingdom; MRC Unit for Lifelong Health and Ageing, UCL, Floor 5, 1-19 Torrington Place, London, United Kingdom; William Harvey Research Institute, Barts C The London Faculty of Medicine C Dentistry, Queen Mary University of London, Charterhouse Square, London EC1 M 6BQ, UK; Barts Interventional Group, Barts Heart Centre, Barts Health NHS Trust, London, UK; Queen Mary University of London, 27 Mile End Rd, Bethnal Green, London, United Kingdom; Institute of Cardiovascular Science, Faculty of Population Health, University College London, 69-75 Chenies Mews, London WC1E 6HX, United Kingdom. 2UCL British Heart Foundation Center of Excellence, 69-75 Chenies Mews, London WC1E 6HX, United Kingdom. 3Department of Cardiology, Amsterdam Cardiovascular Sciences, Amsterdam University Medical Centres, University of Amsterdam, Amsterdam UMC, locatie AMC Postbus 22660, 1100 DD Amsterdam Zuidoost, the Netherlands

## Abstract

**Background:** Premature, or early-onset, coronary artery disease (CAD) carries lifelong consequences. Ethnic inequalities in CAD are established in the UK, but risk estimates mostly derive from events after middle age, while ethnicity is aggregated for South Asian and Black populations. Disaggregated, sex-specific risk estimates would permit better recognition and more targeted prevention.

**Methods:** We used Clinical Practice Research Datalink (CPRD) Aurum, an English primary care database linked to hospital, mortality, and deprivation data. We included adults aged 18–45 of White European, South Asian (Indian, Pakistani, Bangladeshi), or Black (African, Caribbean) ethnicity, followed for up to 20 years. Incident premature CAD (onset ≤45) was a first myocardial infarction or coronary revascularisation. We estimated age-adjusted and fully adjusted incidence rate ratios (IRRs) versus White Europeans by Poisson regression, testing an ethnicity– sex interaction.

**Findings:** Among 14.8 million adults contributing 80.9 million person-years, 16,001 premature CAD events occurred (77.6% in men). In men, the combined South Asian age-adjusted IRR was 1.87 [95% CI 1.77, 1.99], ranging from 1.39 [1.28, 1.50] in Indian to 2.79 [2.52, 3.07] in Bangladeshi men, persisting after full adjustment and already evident at ages 18-26. The combined Black IRR was 0.64 [0.57, 0.71], lowest in African (0.57[0.50, 0.65]) and highest in Caribbean men (0.82 [0.68, 0.98]). In women, the combined South Asian IRR showed no overall excess (1.10 [0.95, 1.26]), concealing a clear excess in Pakistani women (1.56 [1.29, 1.89]). The ethnicity–sex interaction was significant (p<0.001); the male-to-female ratio was highest in Bangladeshi individuals (8.4:1 versus 3.3:1 in White Europeans).

**Interpretation:** Aggregated ethnic categories conceal sex-specific subgroups at high risk of premature CAD, a risk already present in early adulthood. Current screening and health check programmes beginning at age 40, start too late to reach these higher-risk, underserved groups.

**Funding:** Kusuma Trust and NIHR UCLH BRC.

**Research in context:** We searched MEDLINE (via PubMed) from database inception to 27 July 2026, restricted to English-language articles. We combined subject headings and free-text terms for coronary artery disease, myocardial infarction, and ischaemic heart disease with terms for premature or early-onset disease and terms for ethnicity, race, and specific ethnic groups. The full search strategy is provided in the appendix. It is well established that South Asian populations in the UK and internationally develop coronary disease earlier and at higher rates than White populations. This evidence comes mainly from hospital case series, cross-sectional risk-factor surveys, and mortality data, and uses aggregated ethnic categories such as “South Asian” and “Black”. The few studies that disaggregated these categories, found substantial within-group variation, but no study has estimated premature coronary artery disease incidence across disaggregated ethnic subgroups in England using linked primary care, hospital, and mortality records, and no study has examined how sex modifies ethnic differences within those disaggregated groups.

**Added value of this study:** In 14.8 million adults aged 18–45 years, the largest UK study of premature CAD to date, we show that ethnic inequalities are already established in early adulthood, within an age band existing risk screening does not reach. Disaggregation reveals heterogeneity that aggregated analyses obscure: Bangladeshi men carry nearly threefold and Pakistani men approximately twofold the coronary risk of White European men, with a smaller but consistent excess in Indian men, while African incidence sits well below the White rate and Caribbean approaches it. A significant ethnicity–sex interaction exposes a clear excess in Pakistani women entirely hidden within a null combined South Asian female estimate, identifying a high-risk group not previously described.

**Implications of all the available evidence:**

- Disaggregated, sex-specific ethnic reporting should be standard practice across CVD and broader clinical research. Aggregated categories obscure the groups at highest risk and can actively misinform prevention, as here, where a null result for South Asian women overall concealed a clear excess in Pakistani women.
- The elevated risk is established in early adulthood years before the National Health Service (NHS) health check at age 40, therefore current programme reaches too late. Earlier, targeted CVD risk assessment in Bangladesh, Pakistani men and Pakistani women, particularly in deprived communities is warranted.

## Introduction

Coronary artery disease (CAD) is the leading cause of premature cardiovascular death in the United Kingdom (UK).^(1)^ Early onset CAD or ‘premature’ CAD, is variably defined, but increasingly recognised as onset at or before age 45,^(2)^ and carries a lifelong burden: decades of secondary prevention and complications, recurrent admissions, lost working years, and premature death. Despite improvements in cardiovascular disease prevention, premature CAD incidence and mortality has risen in several high-income countries^(3–6)^.

While ethnic differences in cardiovascular disease are well established ^(7,8)^ they are largely described for all ages combined, overlooking potential excess vulnerability at younger ages.^(9)^ Furthermore, most analyses combine multiple minority ethnic groups into a single entity, despite the multitude of ancestral, cultural and socioeconomic sub-differences that exist within them. Such aggregation can obscure minorities at higher risk ^(10)^ and produce risk estimates that may misinform clinical decisions,^(11)^ not least because those estimates inform CVD risk-estimation tools that guide prevention.

Sex adds a further layer to this challenge. Women develop CAD later than men, so premature CAD risk naturally dominates in men, but affected women who do present early are an under recognised, very high risk group.^(12)^ Importantly, key drivers for premature CAD risk in women are themselves linked to ethnicity – for example diabetes confers a threefold excess risk of CAD in women^(13)^ versus two-fold excess in men and is more prevalent in South Asian populations.^(14)^ As such, female risk for premature CAD may be disproportionate across ethnicities and their disaggregated subgroups. How sex interacts with ethnicity in premature CAD is unclear.

To address these questions, we used linked primary and secondary care data on more than 21 million adults in England, followed for up to 20 years. We focused on the White European, South Asian, and Black populations, the largest ethnic groups in the UK^(15)^, disaggregating each into its major constituent subgroups. We aimed to (i) quantify ethnic and sex-specific differences in premature CAD incidence across South Asian (Indian, Pakistani, Bangladeshi) and Black (African, Caribbean) subgroups; (ii) characterise their cardiometabolic and socioeconomic risk factor profiles; (iii) determine to what extent these risk factors explain the differences among ethnic groups.

## Methods

### Data sources and study population

We used anonymised primary care electronic health records from the Clinical Practice Research Datalink (CPRD) Aurum (March 2024 build), a validated longitudinal database representative of the UK population^(16)^ from 1995 to March 2024.^(17)^ It covers ∼ 47 million patients from 1,596 English general practices using EMIS software.

Records were linked at the individual level to Hospital Episode Statistics (HES) admitted patient care data, patient-level Index of Multiple Deprivation (IMD), and Office for National Statistics (ONS) death registration data (linkage sources and coverage periods in table S16). We included CAD free adults aged ≥18 years registered with a CPRD Aurum practice for at least six months between 1 April 2004 and 1 March 2024, whose records met CPRD quality standards and were approved for HES and ONS linkage. Follow-up began at the latest of practice up-to-standard date, six months after current registration, or study start, and was censored at the earliest of first CAD event, death, deregistration from current CPRD practice, last practice collection date, or study end. Events identified in HES were included only if they occurred during active CPRD registration.

### Exposure: ethnicity

Ethnicity was ascertained from SNOMED CT codes in CPRD Aurum, supplemented from HES records where primary care data were missing, an approach shown to maximise completeness.^(18)^ Ethnicity was coded into disaggregated South Asian subgroups according to geographical ancestry (Indian, Pakistani, Bangladeshi), disaggregated Black subgroups (Caribbean and African), White European (reference), Chinese, Other Asian, Mixed, Other Black and Other. Individuals with missing ethnicity were retained as a separate indicator category (supplementary table S1).

### Outcome

The primary outcome was incident CAD, defined as the first recorded composite of myocardial infarction (MI) or coronary revascularisation in primary care (SNOMED CT, EMIS local codes) or secondary care (ICD-10 for diagnoses, OPCS-4 for revascularisation). A strict incident definition required at least six months of prior CPRD registration with no CAD record; any CAD recorded within this washout was classified as prevalent and excluded. Code lists followed best practice guidance^(19)^, were cross-referenced against CALIBER^(20)^ and the London School of Hygiene and Tropical Medicine (LSHTM) phenotype library^(21)^, and were independently reviewed by two cardiologists.

### Covariates

Covariates were based on the closest coded record on or before the index date (date of study entry). Code lists for covariate definitions were obtained from the University of Exeter CPRD code list repository^(22)^. Smoking status was categorised as current, ex, never, or missing, using the most recent recorded status. Hypertension, hyperlipidaemia, family history of premature CAD and diabetes mellitus were each defined by the presence of relevant diagnostic codes or disease-specific prescribing, with absence of a code interpreted as absence of the diagnosis. Consequently, missing data were only possible for body mass index (BMI) and smoking, handled with a separate “missing” category. BMI was derived from the closest recorded measurement before the index date and categorised using standard WHO thresholds and categorised as underweight (<18.5), normal (18.5-24.9), overweight (25-29.9) and Obese (30+). BMI was missing for 30.2% of individuals and smoking status for 17.7%. Socioeconomic deprivation was measured using Index of Multiple Deprivation (IMD) quintile, from patient-level postcode linkage where available (99.9%).

### Statistical analysis

Baseline characteristics were summarised by ethnicity and sex. Premature CAD incidence rates per 1000 person-years were calculated by sex and ethnic sub-group. Cumulative incidence of premature CAD was then plotted overall and by ethnic subgroup and sex.

We estimated incidence rate ratios (IRRs) by ethnic group for premature CAD (ages 18–45) using Poisson regression with robust standard errors to account for clustering by primary care practice. Person-time was portioned by attained age using Lexis expansion, with White European individuals as the reference group. We examined an ethnicity–sex interaction and stratified analyses by sex. To ascertain whether ethnic differences varied by age, IRRs were additionally estimated within tertilised age bands (18–26, 27–35, 36–45) by ethnicity and sex.

Within each sex, we assessed risk factor contributions in two ways. an individual-covariate analysis: age-adjusted (Model 1,) then each covariate added singly (IMD, diabetes, smoking, hypertension, hyperlipidaemia, family history, and BMI; Models 2–8), alongside a fully adjusted model including all covariates (Table S5&6). Second, a cumulative addition analysis, adding covariates in the same order (Table S7&8). The age-adjusted estimate was the primary measure of the ethnic disparity, and the fully adjusted estimate the residual after all measured factors.

To estimate the population-level burden, we applied the White European premature CAD incidence rate, together with the age-adjusted IRR and fully adjusted IRR for each ethnicity–sex group, to the Census 2021 population of England and Wales aged 18–45,(23) deriving the expected excess (or fewer) events over 5 years; this is an illustrative estimate rather than a precise projection.

We performed several sensitivity analyses. The **main analysis was a complete-case analysis** restricted to individuals with recorded ethnicity (89% of those eligible): missing BMI and smoking were handled as above. Complete-case analysis and multiple imputation are not systematically ordered with respect to bias, and complete-case analysis remains valid under plausible assumptions in this setting. ^(24)^ Analyses were repeated excluding those with missing BMI or missing ethnicity and, alternatively, treating missing BMI and missing ethnicity as separate categories. Each comparison was also repeated in datasets restricted to White European individuals and the relevant ethnic group. Analyses were conducted in R (version 4.4.1).

## Results

### Cohort

The full CPRD Aurum cohort comprised 21,495,742 adults, of whom 14,800,928 aged 18–45 were eligible (appendix, cohort flowchart Figure S6). These contributed 80,868,144 person-years, during which 16,001 incident premature CAD events were recorded (12,411 [77.6%] male; 3,590 [22.4%] female). White European, South Asian and Black individuals together comprised 11,606,321 (78.4%) and formed the main comparisons (Table 1); Indian and African were the largest subgroups within South Asian and Black categories respectively. Characteristics of other ethnic groups are reported in the appendix (Table S1). Ethnicity was missing for 1,704,558 (11.5%), who were excluded from the main analyses (Table S1).

**Table 1:** Baseline characteristics of the cohort (aged 18-45 years), by sex and ethnicity † Combined columns show the aggregated ethnic group; subgroup columns disaggregate it. Mean (SD) for continuous variables. BMI, body mass index; IMD, Index of Multiple Deprivation.

|  | South Asian |  |  |  |  | Black |  |  |
| --- | --- | --- | --- | --- | --- | --- | --- | --- |
|  | White | Combined† | Indian | Pakistani | Bangladeshi | Combined† | African | Caribbean |
| **Male** |  |  |  |  |  |  |  |  |
| N | 4,757,930 | 465,247 | 245,212 | 151,229 | 68,806 | 296,401 | 226,805 | 69,596 |
| Age, yrs | 29.3 (8.7) | 29.0 (8.0) | 29.7 (7.7) | 28.2 (8.1) | 28.6 (8.3) | 30.2 (8.8) | 30.2 (8.6) | 30.3 (9.2) |
| IMD most deprived quintile, N (%) | 903,365 (19.0) | 130,867 (28.1) | 44,339 (18.1) | 57,267 (37.9) | 29,261 (42.5) | 114,495 (38.6) | 88,775 (39.1) | 25,720 (37.0) |
| Diabetes mellitus, N (%) | 58,203 (1.2) | 11,691 (2.5) | 6,078 (2.5) | 3,434 (2.3) | 2,179 (3.2) | 6,003 (2.0) | 4,616 (2.0) | 1,387 (2.0) |
| Hypertension, N (%) | 96,955 (2.0) | 11,578 (2.5) | 6,544 (2.7) | 3,161 (2.1) | 1,873 (2.7) | 10,678 (3.6) | 8,289 (3.7) | 2,389 (3.4) |
| Hyperlipidaemia, N (%) | 68,821 (1.4) | 14,431 (3.1) | 7,139 (2.9) | 4,213 (2.8) | 3,079 (4.5) | 4,956 (1.7) | 3,942 (1.7) | 1,014 (1.5) |
| Current smoker, N (%) | 1,313,220 (27.6) | 106,476 (22.9) | 46,695 (19.0) | 38,114 (25.2) | 21,667 (31.5) | 65,703 (22.2) | 43,842 (19.3) | 21,861 (31.4) |
| Obesity (≥30 kg/m<sup>2</sup>), N (%) | 414,312 (8.7) | 35,181 (7.6) | 18,150 (7.4) | 13,364 (8.8) | 3,667 (5.3) | 29,367 (9.9) | 21,788 (9.6) | 7,579 (10.9) |
| Family history, N (%) | 379,997 (8.0) | 46,840 (10.1) | 21,838 (8.9) | 17,002 (11.2) | 8,000 (11.6) | 10,366 (3.5) | 6,897 (3.0) | 3,469 (5.0) |
| **Female** |  |  |  |  |  |  |  |  |
| N | 5,324,626 | 425,452 | 233,084 | 132,141 | 60,227 | 336,288 | 254,939 | 81,349 |
| Age, yrs | 28.7 (8.2) | 28.4 (7.6) | 29.2 (7.4) | 27.7 (7.9) | 27.1 (7.6) | 29.5 (8.2) | 29.4 (8.0) | 29.9 (8.7) |
| IMD most deprived quintile, N (%) | 969,285 (18.2) | 117,427 (27.6) | 39,104 (16.8) | 51,867 (39.3) | 26,456 (43.9) | 127,768 (38) | 97,488 (38.2) | 30,280 (37.2) |
| Diabetes mellitus, N (%) | 79,385 (1.5) | 13,321 (3.1) | 5,995 (2.6) | 4,547 (3.4) | 2,779 (4.6) | 7,096 (2.1) | 5,056 (2.0) | 2,040 (2.5) |
| Hypertension, N (%) | 107,399 (2.0) | 8,503 (2.0) | 4,006 (1.7) | 2,658 (2.0) | 1,839 (3.1) | 13,988 (4.2) | 9,779 (3.8) | 4,209 (5.2) |
| Hyperlipidaemia, N (%) | 40,262 (0.8) | 5,562 (1.3) | 2,351 (1.0) | 2,386 (1.3) | 1,459 (2.4) | 3,354 (1.0) | 2,386 (0.9) | 968 (1.2) |
| Current smoker, N (%) | 1,375,504 (25.8) | 40,985 (9.6) | 14,630 (8.5) | 13,120 (9.9) | 6,119 (10.2) | 43,071 (12.8) | 25,130 (9.9) | 17,941 (22.1) |
| Obesity (≥30 kg/m<sup>2</sup>), N (%) | 594,518 (11.2) | 42,050 (9.9) | 19,228 (8.2) | 16,802 (12.7) | 6,020 (10.0) | 64,278 (19.1) | 48,014 (18.8) | 16,264 (20.0) |
| Family history, N (%) | 513,009 (9.6) | 46,893 (11.0) | 22,985 (9.9) | 16,234 (12.3) | 7,674 (12.7) | 15,091 (4.5) | 9,600 (3.8) | 5,491 (6.7) |

Baseline characteristics are shown in Table 1. Age at entry was similar across ethnic groups and sexes. Deprivation differed markedly by ethnicity but little by sex; within the South Asian category, Pakistani and Bangladeshi men were far more likely than Indian men to live in the most deprived quintile (37.9% and 42.5% vs 18.1%). Diabetes was twofold more prevalent in South Asian than White groups, and around 1.5-fold higher in Black groups. Hypertension was higher across almost all Black subgroups and smoking was highest in Bangladeshi and Caribbean men (both ∼ 31%).

### Cumulative incidence to age 45

Cumulative incidence of CAD diverged by ethnicity from around age 35 years in both sexes: South Asian groups had the highest incidence at 45 years, White Europeans intermediate, and Black groups lowest (Figure 1). In men, cumulative incidence at 45 years was 1.42% [95% CI 1.35, 1.50] in South Asian men overall, ranging from 0.98% [0.90, 1.07] in Indian to 2.20% [1.98, 2.45] in Bangladeshi men – over three times the White European rate of 0.70% [0.69, 0.72].

**Figure 1:**
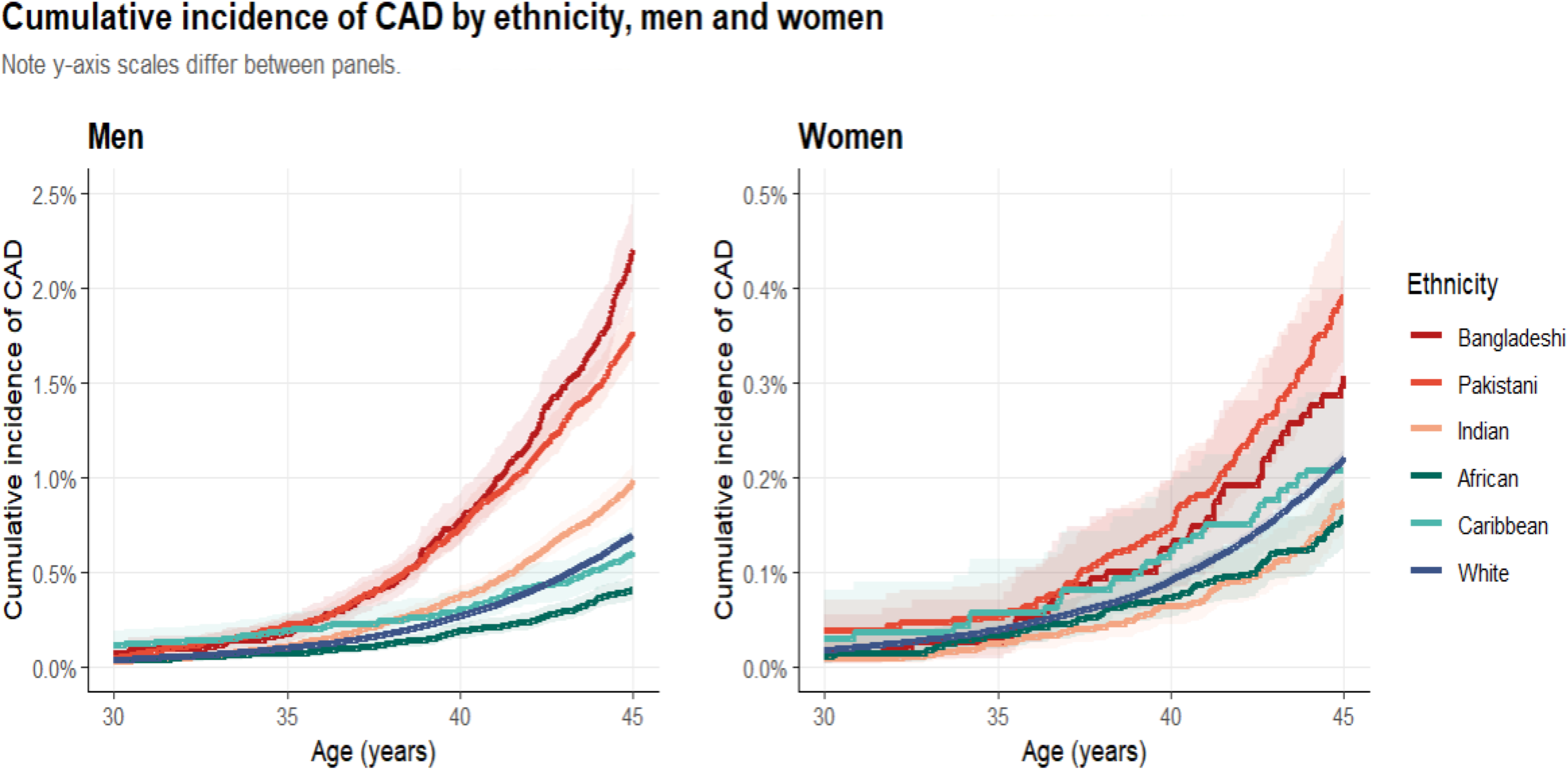
Cumulative incidence of coronary artery disease from 30 till 45 years, by disaggregated ethnic group and sex. Cumulative incidence estimates were derived from Kaplan-Meier survival functions (1 − S(t)). Shaded ribbons represent 95% confidence intervals based on the log-log transformation. Note that y-axis scales differ between panels to accommodate the lower absolute incidence in women; relative differences between ethnic groups should be compared within, not across, panels. Abbreviations: CAD, coronary artery disease.

In women, the South Asian estimate (0.26% [0.23, 0.30]) resembled White European women (0.22% [0.21, 0.23]) overall, concealing a higher incidence in Pakistani women 0.39% [0.32, 0.47]. Black cumulative incidence was at or below the White rate in both sexes, closest in Caribbean men (0.60% [0.49, 0.74]) and lowest in African men (0.41% [0.36, 0.47]).

Unadjusted incidence rates varied substantially by ethnicity and sex (Table 2). In men, rates varied more than twofold within South Asian category (Indian 0.46 [0.42,0.50] to Bangladeshi 0.92 [0.83,1.01] per 1000 person years) and were lower in Black men. In women, ethnic differences were smaller, but Pakistani rate (0.16 [0.13, 0.19]) was markedly higher than in Indian or Bangladeshi women.

**Table 2.**
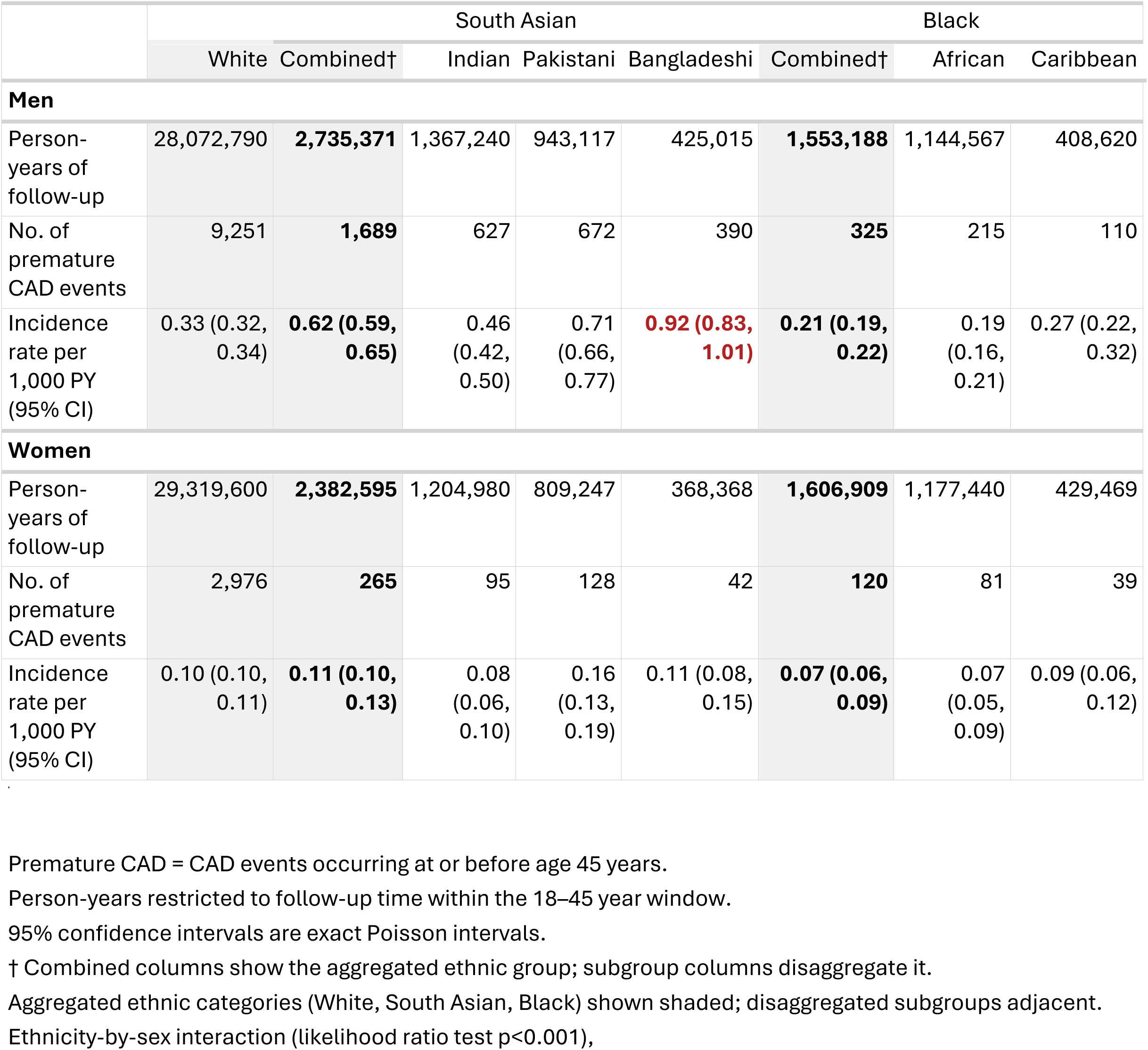
Incidence of premature CAD (ages 18–45 years), by sex and ethnicity Premature CAD = CAD events occurring at or before age 45 years.

There was strong evidence of an ethnicity-by-sex interaction (likelihood ratio test p<0.001): the male to female rate was approximately 3.0 in White and Black groups, 5.8 in Indian, 4.4 in Pakistani and 8.4 in Bangladeshi individuals.

### Risk factor profile

Among individuals with premature CAD, risk factor profiles differed by ethnicity and sex, with the greatest cardiometabolic burden in South Asian subgroups (Figure 2). Within every ethnic group, women with premature CAD carried a higher burden than men, most so in Pakistani and Bangladeshi women, though case numbers were small. Smoking diverged: high in Bangladeshi men (53.6%) and White women (56.9%, exceeding White men), much lower in South Asian (15.1%) and Black (22.5%) women. Deprivation was higher among cases in women than men, varied by ethnicity but not sex.

**Figure 2:**
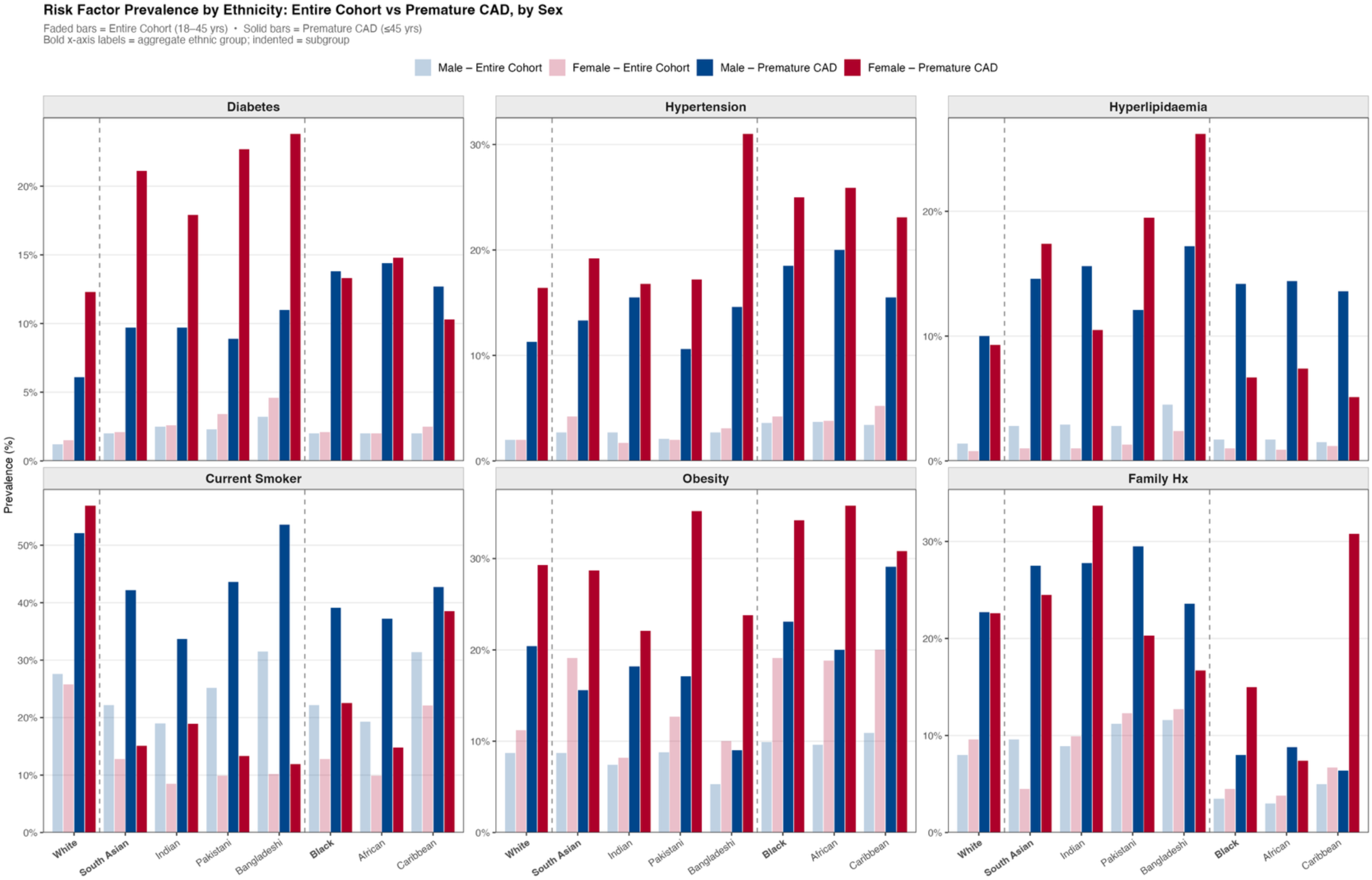
Risk factor profiles by sex, ethnicity and presence of premature CAD. Each panel shows the prevalence of one risk factor: diabetes, hypertension, hyperlipidaemia, current smoking, obesity, and family history of premature cardiovascular disease. Bars show prevalence (%) within each ethnicity, sex, and cohort group. Faded bars denote the entire cohort (adults aged 18 to 45 years). Solid bars denote premature CAD cases (first CAD event at age 45 years or younger). Blue bars denote men. Red bars denote women. Aggregate ethnic groups are shown in bold on the x-axis (White, South Asian, Black). Subgroups are indented (Indian, Pakistani, and Bangladeshi within South Asian; African and Caribbean within Black). Dashed vertical lines separate each aggregate group from its subgroups.

### Ethnic differences in premature CAD rates and potential drivers

Ethnic differences were examined in sex-stratified models (Figure 3, Table S5-8). Among men, the combined South Asian age-adjusted IRR versus White men was 1.87 [95% CI 1.77, 1.99], with subgroup estimates ranging more than two-fold, from 1.39 [1.28, 1.50] in Indian to 2.79 [2.52, 3.07] in Bangladeshi men (Pakistani 2.16 [2.00, 2.33]). The combined Black IRR was 0.64 [0.57, 0.71], lowest in African men (0.57 [0.50, 0.65]) and closest to White in Caribbean men (0.82 [0.68, 0.98]). The higher estimates in South Asians attenuated only modestly after full adjustment (combined 1.75 [1.65, 1.86]; Indian 1.45 [1.33, 1.57]; Pakistani 1.91 [1.75, 2.07]; Bangladeshi 2.21 [1.97, 2.46]), and Black estimates changed little (combined 0.65[0.59, 0.72]).

**Figure 3.**
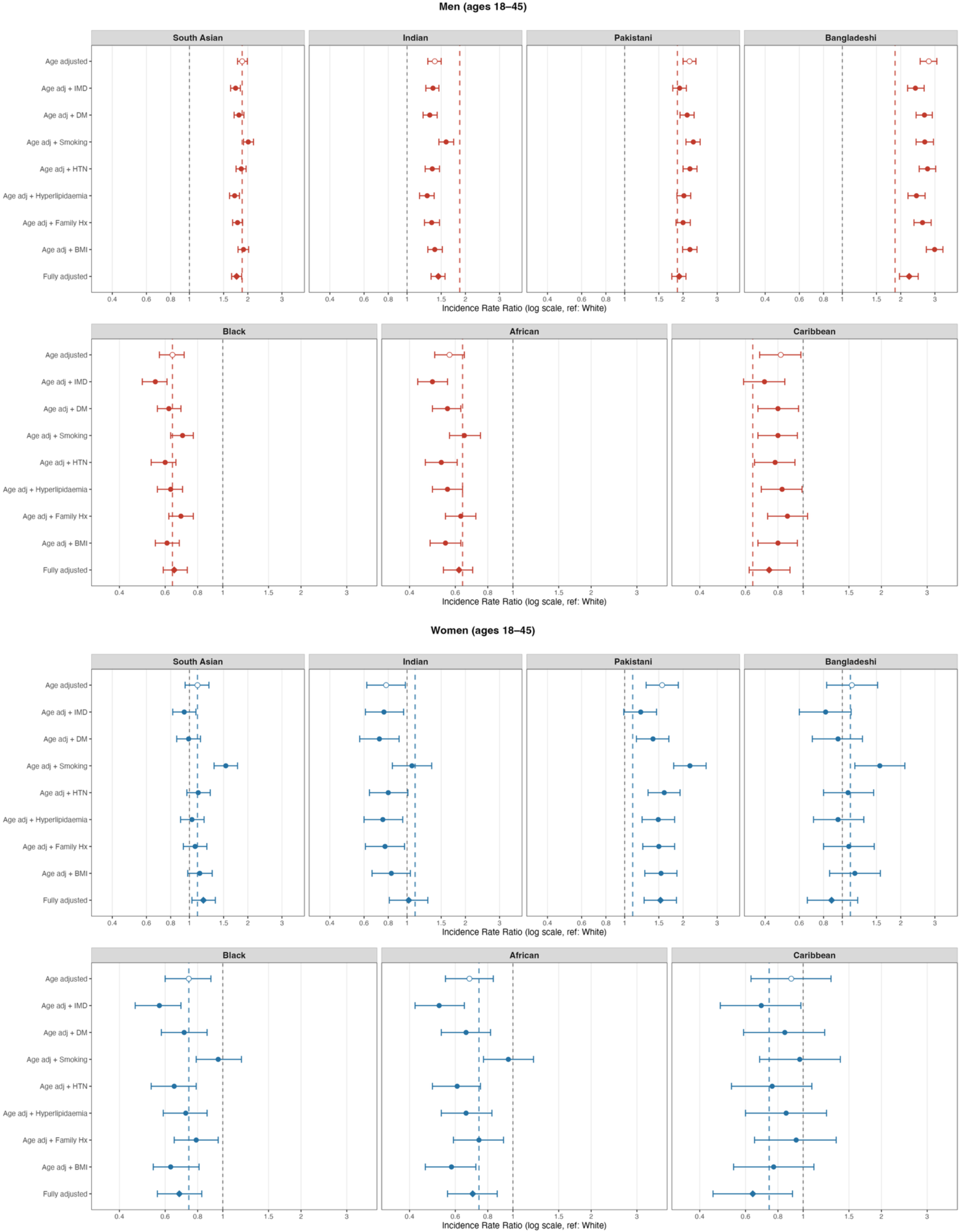
Incidence rate ratios for premature coronary artery disease (age 18–45) by ethnic group and sex, derived from sequentially adjusted Poisson regression models (Models 1–9). Reference: White European. Open circles denote the age-adjusted model (Model 1); diamonds denote the fully adjusted model (Model 9); filled circles denote intermediate models. The coloured vertical dashed line in each subgroup panel represents the age-adjusted IRR of the corresponding aggregate group (South Asian combined or Black combined), carried across subgroup panels to illustrate how aggregate estimates compare with within-group variation. Shaded panels denote aggregate groups. IMD, Index of Multiple Deprivation; BMI, body mass index.

Age-stratified analysis showed this excess is present from early adulthood and follows divergent trajectories by subgroup (Table S12–13). In South Asian men overall, the fully adjusted IRR was 1.79 [1.51, 2.12] at age 18–26 and remained essentially unchanged at 36–45. Disaggregation showed the Bangladeshi excess widening with age, from an age-adjusted IRR of 2.52 [1.84, 3.44] at 18–26 to 3.40 [2.96, 3.89] at 36–45, whereas the Pakistani excess was already 2.5-fold at 18–26 and sustained thereafter. The same early-onset pattern was seen in Pakistani women, with an IRR of 1.65 [1.09, 2.51] at 18–26.

Of the individual covariates, hyperlipidaemia, IMD, and diabetes produced the largest single attenuations in South Asian men (Figure 3, Table S5); in African men, smoking produced the largest attenuation toward the null (0.57 to 0.65), with smaller effects in Caribbean men, who had the second highest smoking prevalence of any group, behind Bangladeshi men.

Among women, the combined South Asian IRR showed no overall excess (1.10 [0.95, 1.26]), yet within it Pakistani women had a clear excess (1.56 [1.29, 1.89]) while Indian (0.78 [0.62, 0.98]) and Bangladeshi (1.12 [0.83, 1.52]) women showed lower or no difference. The combined Black IRR was 0.74 [0.60, 0.90] (African 0.68 [0.55, 0.84]; Caribbean 0.90 [0.63, 1.28]). After full adjustment, only Pakistani women retained an excess (1.53 [1.26, 1.85]); estimates for Indian (1.02 [0.81, 1.28]), Bangladeshi (0.88 [0.66, 1.20]), African (0.70 [0.56, 0.87]), and Caribbean (0.64 [0.45, 0.91]) women were consistent with no difference or lower risk. In Pakistani women, IMD, diabetes, and hyperlipidaemia produced the largest single attenuations (Figure 3, Table S6).

### Population-level burden

Applied to the 2021 Census population aged 18–45, the age-adjusted disparity corresponded to an estimated 1,389 excess events over 5 years in South Asian men (680 in Pakistani, 431 in Bangladeshi, and 278 in Indian men Table S11). Among women, only Pakistani women showed a clear excess (104 events). All Black subgroups had fewer events than the White comparator throughout, largest deficit in African men (221 fewer).

### Sensitivity analysis

Restricting analyses to White European individuals and each ethnic group did not alter age-adjusted or fully adjusted IRRs in either sex (Table S4&5). Including missing ethnicity as a separate category did not change IRRs or the ethnicity–sex interaction, indicating their exclusion from the main analysis did not bias estimates.^(24)^ Restriction to individuals with recorded BMI produced modestly higher fully adjusted estimates in Pakistani and Bangladeshi men and women, with Indian and Black estimates unchanged, suggesting conservative estimates in Pakistani and Bangladeshi groups. Including fatal CAD cases identified only from death registration, absent from primary care and hospital data, did not change IRRs (table S13 and S14).

## Discussion

In a national cohort of 14.8 million adults aged 18–45, premature CAD incidence was elevated in specific ethnic subgroups, and the pattern differed by sex in ways that aggregate categories obscured. The South Asian male excess was driven by Bangladeshi incidence, nearly three times the White European rate (age-adjusted IRR 2.79[2.52, 3.07]). Pakistani men showed a twofold excess and Indian men a smaller but consistent one. Differences persisted after adjustment for measured cardiometabolic and socioeconomic factors. Among women, the aggregate South Asian rate matched the White rate (age-adjusted IRR 1.10 [0.95, 1.26]), yet this comparison hid a clear excess confined to Pakistani women (1.56 [1.29, 1.89]). Black subgroups had lower incidence in every model despite higher hypertension and diabetes prevalence, with Caribbean incidence near the White rate and African incidence well below it.

Prior studies characterised these differences in middle and older age. UK Biobank (UK-B) and the Southall and Brent Revisited (SABRE) study consistently identify elevated CAD risk in South Asians, lower CAD risk in Black individuals, and higher stroke burden in both, but recruited in mid-to-later life and cannot establish when the disparities begin. ^(9,25)^. In the BCIS PCI registry, South Asians presented younger than White Europeans, but at a mean age near 60.^(8)^ We show the disparity is well established in early adulthood, years before events accrue, identifying a key window for prevention. To our knowledge, this is the first UK cohort large enough to estimate premature CAD incidence by ethnic subgroup and sex simultaneously, exposing the ethnicity-by-sex interaction and the hidden excess in Pakistani women.

Much of the South Asian male excess persisted after adjustment. Hyperlipidaemia, deprivation, and diabetes drove the largest attenuations, but a clear residual remained, and conventional risk factors explained only a modest share of incidence, equally across ethnic groups. Plausible factors may include higher lipoprotein(a), ectopic adiposity, duration of metabolic dysfunction, including insulin resistance before overt diabetes, and polygenic risk. The Indian excess persisted, and even widened, on adjustment despite deprivation comparable to White Europeans, highlighting that measured factors do not account for it.

In women, only Pakistani women showed a clear excess. Deprivation and diabetes attenuated much of it, exposures recorded years before the first event, so the pathway is modifiable and identifiable in advance. In White women, smoking was the dominant contributor (56.9%).In South Asian women, the profile was metabolic, not behavioural. Diabetes (21% vs 12%) and hyperlipidaemia (17% vs 9%) were both raised, despite smoking rates less than a third of White women (15% vs 57%). Diabetes carries a larger excess CVD risk in women than in men, and since smoking is far lower here, their true metabolic susceptibility may exceed the observed excess.

Lower coronary incidence in Black adults survived adjustment, but the combined estimate hid a contrast. African adults had the lowest rates of any group; Caribbean men approached the White rate despite high smoking, developing less CAD than predicted by their risk-factor profile. This fits a cardiovascular phenotype in which hypertensive heart disease, heart failure, and stroke predominate over coronary atherosclerosis, as in SABRE. ^(26,27)^. Why African rates fall below Caribbean is unclear. One possibility is a healthy migrant effect, since African migration to the UK is more recent, but CPRD holds no migration data, and known migration patterns would explain only part of the gap. ^(28)^

The principal strengths are scale and granularity: with 14.8 million adults and up to 20 years of follow-up, this is far larger than any previous study of premature CAD and permitted disaggregated estimation with narrow confidence intervals where events were sparse.

Several limitations apply. Ethnicity data were ∼90% complete, though sensitivity analyses addressing this were reassuring; missing BMI (30.2%) and smoking (17.7%) were handled with indicator categories, which can leave residual confounding, though restriction to recorded BMI suggested our South Asian estimates are conservative (Table S3&4). Distinguishing true differences in incidence from differences in diagnostic practice is inherent to routine data, but two features limit its effect here. Our primary outcome, MI or coronary revascularisation, comprises hard clinical events less susceptible to differential coding than symptom-based diagnoses. These were ascertained across linked primary care, hospital, and mortality records, so an event missed in one source is captured in another. Comorbidities, defined from codes and prescribing, remain subject to differential recording, and residual confounding from unmeasured factors cannot be excluded. Family history relies on it being elicited and coded, so it is likely under recorded, though this should apply equally across all ethnic groups.

Across both sexes, the disparity is established by the mid-thirties, years before the NHS Health Check begins at 40, so the window for prevention opens well before these patients reach current clinical pathways.

Current tools and thresholds miss this window. Treatment thresholds rest on 10-year absolute risk, such that a young adult with high relative risk can still fall below them. A 38-year-old Bangladeshi man with hypertension carries over three times the risk of a White European man of the same age, yet, in the absence of other risk factors, remains below the 10% statin threshold according to QRISK3, the most commonly used CVD risk scoring system in the UK. Existing tools also underestimate risk in these groups even where ethnicity is a factor in prediction models. In UK Biobank observed risk in South Asians exceeded QRISK3 predictions despite its ethnicity-specific adjustment;^(25)^ in the SABRE study, QRISK2 captured only half the observed risk in South Asian women.^(29)^

Four implications follow. First, screening and public awareness should begin well before 40 in Bangladeshi and Pakistani men and Pakistani women, particularly in deprived communities. Risk scoring tools should be recalibrated using larger, more representative contemporary datasets for the under-45s using disaggregated groups. Second, in acute settings, heightened clinical suspicion of MI is warranted when managing young South Asian men and Pakistani women with chest pain. Third, prevention in Pakistani women in particular should prioritise earlier and preclinical diagnoses of diabetes, hyperlipidaemia, and raised blood-pressure. Fourth, low premature CAD rates in African and Caribbean adults should not be mistaken for low cardiovascular risk overall; reported stroke and heart failure burden means blood-pressure and diabetes screening should stay a priority. Across all these groups premature CAD strikes in the most economically active years, at the youngest and costliest end of a burden (80,000 working years and £3 billion in lost productivity annually),^(30)^ making early prevention both a clinical and economic priority.

Disaggregating commonly used ethnic groupings reveals hidden sex specific risks for Bangladeshi men and Pakistani men and women. For these high-risk groups, risk assessments before the age of 40 could represent important CVD prevention opportunities that will also address persisting ethnic health disparities. In Black adults, low CAD risk should not be mistaken for low CVD risk.

## Declarations

### Ethical approval

CPRD holds annual ethical approval from the UK’s Health Research Authority (HRA) Research Ethics Committee (REC) (East Midlands – Derby, REC reference 05/MRE04/87), under which individual observational studies do not require additional ethical approval. Data used in this study was provided under protocol number 23_003525 (CPRD Aurum, March 2024 release).

### Data sharing

CPRD Aurum data are available via approved application to CPRD. Code lists will be made available in a public repository (Github) on publication.

### Patient and public involvement

Fifteen individuals with lived experience of premature coronary artery disease, including ten of South Asian ethnicity, were consulted before the study and confirmed the relevance and priority of the research question. Patients were not otherwise involved in the design, conduct, analysis, or reporting of the study. Contact details were not collected, so findings could not be disseminated back to those involved.

### Contributors

NN, SE, NC, and RSP conceived and designed the study and developed the statistical analysis plan. NN did the data curation and statistical analysis and wrote the first draft of the manuscript. SP contributed to the development of the code lists and data queries. JE contributed to data curation and resolved data quality issues. SE, NC, and RSP supervised the study. SP, JE, SQ, SD, KR, FC, AJ, AFS, and ADH critically reviewed the manuscript for important intellectual content. All authors reviewed and approved the final version of the manuscript and agree to be accountable for all aspects of the work. NN and RSP were responsible for the decision to submit for publication.

## Data Availability

All data produced in the present study are available upon reasonable request to the authors
All data produced in the present work are contained in the manuscript

## References

1. Ischaemic heart diseases deaths including comorbidities, England and Wales – Office for National Statistics [Internet]. [cited 2026 Apr 20]. Available from: https://www.ons.gov.uk/peoplepopulationandcommunity/birthsdeathsandmarriages/deaths/bulletins/ischaemicheartdiseasesdeathsincludingcomorbiditiesenglandandwales/2019registrations

2. Ali-Patel A, Natarajan N, Bijral M, Diamondali S, Joshi A, Rathod K, et al. 7-004 Defining premature myocardial infarction: a scoping review. Heart. 2025 Sep 1;111(Suppl 3):A202–3. doi:10.1136/heartjnl-2025-BCS.200

3. bhf-cvd-statistics-uk-factsheet-jan26.pdf [Internet]. [cited 2026 Apr 17]. Available from: https://www.bhf.org.uk/-/media/files/for-professionals/research/heart-statistics/bhf-cvd-statistics-uk-factsheet-jan26.pdf?rev=26534e1487094dbd806277891baef112

4. Li H, Zheng J, Qian F, Zou X, Zou S, Wu Z, et al. Demographic and regional trends of acute myocardial infarction-related mortality among young adults in the US, 1999–2020. Npj Cardiovasc Health. 2025 Mar 4;2(1):9. doi:10.1038/s44325-025-00046-w

5. Arora S, Stouffer GA, Kucharska-Newton AM, Qamar A, Vaduganathan M, Pandey A, et al. Twenty Year Trends and Sex Differences in Young Adults Hospitalized With Acute Myocardial Infarction. Circulation. 2019 Feb 19;139(8):1047–56. doi:10.1161/CIRCULATIONAHA.118.037137

6. Gabet A, Danchin N, Juillière Y, Olié V. Acute coronary syndrome in women: rising hospitalizations in middle-aged French women, 2004–14. Eur Heart J. 2017 Apr 7;38(14):1060–5. doi:10.1093/eurheartj/ehx097

7. Razieh C, Zaccardi F, Miksza J, Davies MJ, Hansell AL, Khunti K, et al. Differences in the risk of cardiovascular disease across ethnic groups: UK Biobank observational study. Nutr Metab Cardiovasc Dis. 2022 Nov;32(11):2594–602. doi:10.1016/j.numecd.2022.08.002

8. Jones DA, Gallagher S, Rathod KS, Redwood S, de Belder MA, Mathur A, et al. Mortality in South Asians and Caucasians After Percutaneous Coronary Intervention in the United Kingdom: An Observational Cohort Study of 279,256 Patients From the BCIS (British Cardiovascular Intervention Society) National Database. JACC Cardiovasc Interv. 2014 Apr 1;7(4):362–71. doi:10.1016/j.jcin.2013.11.013

9. Tillin T, Hughes AD, Mayet J, Whincup P, Sattar N, Forouhi NG, et al. The relationship between metabolic risk factors and incident cardiovascular disease in Europeans, South Asians, and African Caribbeans: SABRE (Southall and Brent Revisited) –-a prospective population-based study. J Am Coll Cardiol. Apr 30;61(17):1777–86. doi:10.1016/j.jacc.2012.12.046

10. Liang PS, Kwon S, Cho I, Trinh-Shevrin C, Yi S. Disaggregating racial and ethnic data: a step toward diversity, equity, and inclusion. Gastroenterology. 2023 Mar;164(3):320–4. doi:10.1053/j.gastro.2023.01.008 PubMed PMID: 36822735; PubMed Central PMCID: PMC10983115.

11. Gordon NP, Lin TY, Rau J, Lo JC. Aggregation of Asian-American subgroups masks meaningful differences in health and health risks among Asian ethnicities: an electronic health record based cohort study. BMC Public Health. 2019 Nov 25;19:1551. doi:10.1186/s12889-019-7683-3 PubMed PMID: 31760942; PubMed Central PMCID: PMC6876105.

12. Hammond J, Salamonson Y, Davidson P, Everett B, Andrew S. Why do women underestimate the risk of cardiac disease? A literature review. Aust Crit Care. 2007 May 1;20(2):53–9. doi:10.1016/j.aucc.2007.02.001

13. Huxley R, Barzi F, Woodward M. Excess risk of fatal coronary heart disease associated with diabetes in men and women: meta-analysis of 37 prospective cohort studies. BMJ. 2006 Jan 14;332(7533):73–8. doi:10.1136/bmj.38678.389583.7C PubMed PMID: 16371403; PubMed Central PMCID: PMC1326926.

14. Tillin T, Sattar N, Godsland IF, Hughes AD, Chaturvedi N, Forouhi NG. Ethnicity-specific obesity cut-points in the development of Type 2 diabetes – a prospective study including three ethnic groups in the United Kingdom. Diabet Med. 2015 Feb;32(2):226–34. doi:10.1111/dme.12576 PubMed PMID: 25186015; PubMed Central PMCID: PMC4441277.

15. Population of England and Wales [Internet]. 2022 [cited 2026 Jun 8]. Available from: https://www.ethnicity-facts-figures.service.gov.uk/uk-population-by-ethnicity/national-and-regional-populations/population-of-england-and-wales/latest/

16. Wolf A, Dedman D, Campbell J, Booth H, Lunn D, Chapman J, et al. Data resource profile: Clinical Practice Research Datalink (CPRD) Aurum. Int J Epidemiol. 2019 Dec;48(6):1740–1740g. doi:10.1093/ije/dyz034 PubMed PMID: 30859197; PubMed Central PMCID: PMC6929522.

17. Clinical Practice Research Datalink. CPRD Aurum March 2024 [Text/csv]. Clinical Practice Research Datalink; 2024 [cited 2026 Apr 21]. p. Total number of research acceptable patients: 47,413,279, Percentage UK population coverage (current patients only): 16,184,439 of 67,026,300 (24.15%), Median (25th and 75th percentile) follow-up time in years for currently registered patients: 9.55 (3.30-22.22), Patients eligible for linkage: 35,240,366, Total number of GP practices: 1,784, Percentage coverage of UK general practices (currently contributing practices only): 1,596 of 8,036 (19.86%). Available from: https://cprd.com/cprd-aurum-march-2024-dataset doi:10.48329/YXMQ-VK87

18. Mathur R, Bhaskaran K, Chaturvedi N, Leon DA, van Staa T, Grundy E, et al. Completeness and usability of ethnicity data in UK-based primary care and hospital databases. J Public Health Oxf Engl. 2014 Dec;36(4):684–92. doi:10.1093/pubmed/fdt116 PubMed PMID: 24323951; PubMed Central PMCID: PMC4245896.

19. Matthewman J, Andresen K, Suffel A, Lin LY, Schultze A, Tazare J, et al. Checklist and guidance on creating codelists for routinely collected health data research. NIHR Open Res. 2024 Sep 18;4:20. doi:10.3310/nihropenres.13550.2 PubMed PMID: 39345273; PubMed Central PMCID: PMC11437289.

20. Phenotype Library | PH1027/2263 [Internet]. [cited 2026 Apr 20]. Available from: http://phenotypes.healthdatagateway.org/phenotypes/PH1027/version/2263/detail/

21. Bhaskaran K, Matthews A, Smeeth L, Strongman H. Exposure Code List – Cardiovascular disease (ICD-10) [Internet]. London, United Kingdom: London School of Hygiene & Tropical Medicine; 2019 [cited 2026 Apr 20]. Available from: https://datacompass.lshtm.ac.uk/id/eprint/1117/ doi:10.17037/DATA.00001117

22. Exeter-Diabetes/CPRD-Codelists [Internet]. Exeter Diabetes Research Team; 2026 [cited 2026 Jul 27]. Available from: https://github.com/Exeter-Diabetes/CPRD-Codelists

23. UK census data – Office for National Statistics [Internet]. [cited 2026 Apr 20]. Available from: https://www.ons.gov.uk/census/planningforcensus2021/ukcensusdata

24. White IR, Carlin JB. Bias and efficiency of multiple imputation compared with complete-case analysis for missing covariate values. Stat Med. 2010;29(28):2920–31. doi:10.1002/sim.3944

25. Patel AP, Wang M, Kartoun U, Ng K, Khera AV. Quantifying and understanding the higher risk of atherosclerotic cardiovascular disease among South Asians — results from the UK Biobank prospective cohort study. Circulation. 2021 Aug 10;144(6):410–22. doi:10.1161/CIRCULATIONAHA.120.052430 PubMed PMID: 34247495; PubMed Central PMCID: PMC8355171.

26. George J, Mathur R, Shah AD, Pujades-Rodriguez M, Denaxas S, Smeeth L, et al. Ethnicity and the first diagnosis of a wide range of cardiovascular diseases: Associations in a linked electronic health record cohort of 1 million patients. PLOS ONE. 2017 Jun 9;12(6):e0178945. doi:10.1371/journal.pone.0178945

27. M. Talha K, Almas T, Minhas AMK, Salah H, Jamil A, Johnson HM, et al. Disparities in heart failure between White, Black, and Hispanic young adults: insights from the National Health and Nutrition Examination Survey. Ther Adv Cardiovasc Dis. 2024 Mar 24;18:17539447241239814. doi:10.1177/17539447241239814 PubMed PMID: 38523335; PubMed Central PMCID: PMC10962029.

28. Harding S, Rosato M, Teyhan A. Trends for coronary heart disease and stroke mortality among migrants in England and Wales, 1979–2003: slow declines notable for some groups. Heart. 2008 Apr 1;94(4):463–70. doi:10.1136/hrt.2007.122044 PubMed PMID: 17690159.

29. Tillin T, Hughes AD, Whincup P, Mayet J, Sattar N, McKeigue PM, et al. Ethnicity and prediction of cardiovascular disease: performance of QRISK2 and Framingham scores in a UK tri-ethnic prospective cohort study (SABRE—Southall And Brent REvisited). Heart. 2014 Jan 1;100(1):60–7. doi:10.1136/heartjnl-2013-304474

30. Landeiro F, Harris C, Groves D, O’Neill S, Jandu KS, Tacconi EMC, et al. The economic burden of cancer, coronary heart disease, dementia, and stroke in England in 2018, with projection to 2050: an evaluation of two cohort studies. Lancet Healthy Longev. 2024 Aug 1;5(8):e514–23. doi:10.1016/S2666-7568(24)00108-9 PubMed PMID: 39068947.

