## Supplementary material for "Ethnic and sex inequalities in premature coronary artery disease across disaggregated South Asian and Black subgroups in England: a population-based cohort study": Sup

**This appendix forms part of the following article:**

This appendix has been provided by the authors to give readers additional information about their work. It has not been copyedited.

Entire Cohort | N = 21,512,314 | CPRD Aurum

| Ethnicity | N | Age, mean (SD), y | Female, % (n) | IMD least deprived (Q1–Q2), % (n) | IMD most deprived (Q9–Q10), % (n) | Diabetes mellitus, % (n) | Hypertension, % (n) | Hyperlipid-aemia, % (n) | Current smoker, % (n) | Obesity (BMI ≥30), % (n) | Family Hx of CAD, % (n) | CAD events, n | Premature CAD (≤45 y), n | Premature CAD, % of total CAD | Person-years of follow-up |
|---|---|---|---|---|---|---|---|---|---|---|---|---|---|---|---|
| White European | 15,715,535 | 41.4 (19.4) | 52.8% (8,302,685) | 20.7% (3,253,592) | 17.8% (2,795,357) | 4.1% (637,474) | 14.4% (2,257,178) | 9% (1,408,683) | 25.7% (4,044,268) | 13.7% (2,156,638) | 10.8% (1,694,773) | 337,692 | 12,227 | 3.6% | 76,825,025 |
| **South Asian** | **1,090,273** | **34.4 (14.5)** | **48.0% (523,414)** | **9.7% (105,766)** | **27.5% (300,352)** | 7.3% (79,636) | 9.2% (100,787) | 8.1% (88,318) | 16.6% (180,944) | 10.8% (117,995) | **11.3% (123,276)** | **18,118** | **1,954** | **10.8%** | **5,323,618** |
| – Indian | 600,908 | 35.7 (14.8) | 49.1% (294,960) | 13.2% (79,249) | 17.6% (105,641) | 7% (42,355) | 10.1%(60,495) | 8.1% (48,532) | 14.3%(85,680) | 10%(60,370) | 10.5% (62,943) | 9,643 | 722 | 7.5% | 2,790,367 |
| – Pakistani | 338,747 | 33.0 (14.2) | 46.7% (158,315) | 6.1% (20,735) | 38.4% (129,993) | 7.2%(24,418) | 8% (27,156) | 7.6%(25,609) | 18.3% (61,852) | 13.3% (44,906) | 12.3% (41,789) | 6,131 | 800 | 13.0% | 1,758,642 |
| – Bangladeshi | 150,618 | 32.3 (13.7) | 46.6% (70,139) | 3.8% (5,782) | 43.0% (64,718) | 8.5%(12,863) | 8.7%(13,136) | 9.4% (14,177) | 22.2% (33,412) | 8.4% (12,722) | 12.3% (18,544) | 2,344 | 432 | 18.4% | 774,610 |
| **Black** | **816,371** | **36.4 (15.1)** | **52.4% (427,918)** | **4.2% (34,127)** | **38.2% (311,635)** | 6.2% (50,713) | 12.9% (105,607) | 6.5% (52,709) | 17.8% (145,377) | 18.7% (152,350) | **4.7% (38,276)** | **5,324** | **445** | **8.6%** | **3,566,244** |
| – African | 584,372 | 34.4 (13.2) | 52.2% (305,268) | 4.2% (24,765) | 38.6% (225,544) | 4.7%(27,521) | 10% (58,158) | 4.8 %( 28,276) | 14.5% (84,494) | 17.9% (104,504) | 3.9% (22,536) | 2,177 | 296 | 13.6% | 2,445,949 |
| – Caribbean | 231,999 | 41.2 (18.2) | 52.9% (122,650) | 4.0% (9,362) | 37.1% (86,091) | 10% (23,192) | 20.5% (47,449) | 10.5% (24,433) | 26.2% (60,883) | 20.6% (47,846) | 6.8% (15,740) | 3,147 | 149 | 4.7% | 1,120,295 |
| Other Black | 149,033 | 32.9 (13.8) | 50.4% (75,041) | 5.3% (7,958) | 35.5% (52,906) | 3.8% (5,710) | 8.4% (12,506) | 4% (5,990) | 22.4% (33,441) | 15.6% (23,191) | 5.1% (7,618) | 730 | 77 | 10.5% | 673,632 |
| Other Asian | 470,954 | 34.0 (13.6) | 52.4% (246,582) | 11.8% (55,719) | 19.0% (89,454) | 5% (23,424) | 7.2% (34,104) | 6.3% (29,472) | 16.8% (79,122) | 8.7% (40,741) | 8.4% (39,581) | 4,565 | 503 | 11.0% | 2,045,930 |
| Chinese | 313,045 | 28.9 (11.4) | 59.9% (187,498) | 14.1% (44,122) | 15.2% (47,517) | 1.3% (3,996) | 2.5% (7,834) | 1.8% (5,593) | 12.5% (39,071) | 2.7% (8,380) | 2.9% (9,026) | 680 | 42 | 6.2% | 1,149,242 |
| Mixed | 328,954 | 31.2 (13.0) | 53.8% (177,025) | 12.9% (42,328) | 24.7% (81,115) | 2.7% (9,027) | 5.2% (17,259) | 3.4% (11,021) | 24.6% (80,906) | 10.9% (35,746) | 6.9% (22,666) | 1,725 | 193 | 11.2% | 1,431,510 |
| Other | 483,579 | 34.0 (13.7) | 49.9% (241,516) | 9.6% (46,530) | 28.2% (136,323) | 3.1% (14,885) | 5.5% (26,417) | 4.6% (22,315) | 24.1% (116,301) | 11.4% (54,909) | 7.7% (37,341) | 3,864 | 372 | 9.6% | 2,069,196 |
| *Unknown/Missing* | 2,144,570 | 34.9 (16.7) | 36.6% (785,687) | 21.7% (464,534) | 15.5% (331,339) | 1.2% (24,876) | 4.5% (95,917) | 1.8% (38,291) | 18.6% (398,433) | 5.1% (109,618) | 3.5% (74,819) | 4,297 | 188 | 4.4% | 8,960,553 |

*Abbreviations: CAD = Coronary Artery Disease; IMD = Index of Multiple Deprivation; BMI = Body Mass Index; SD = Standard Deviation; Q = Quintile.*
*Premature CAD defined as first CAD event at age ≤45 years. Black aggregate (N = 816,371) comprises African and Caribbean subgroups only; Other Black (N = 149,033) is listed separately. Bold rows = aggregated ethnic groups; subgroups are indented. Unknown/Missing shown last.*

**Table S1. Baseline Characteristics of the Entire Cohort by Ethnicity** entire cohort n= 21,512,314

**Non-Premature CAD Group** | **N = 360,994** | **CPRD Aurum**

| Ethnicity | N | Age, mean (SD), y | Female, % (n) | IMD least deprived (Q1–Q2), % (n) | IMD most deprived (Q9–Q10), % (n) | Diabetes mellitus, % (n) | Hypertension, % (n) | Hyperlipidaemia, % (n) | Current smoker, % (n) | Obesity (BMI ≥30), % (n) | Family Hx of CAD, % (n) | Premature CAD (≤45 y), n | Premature CAD, % of total CAD | Person-years of follow-up |
|---|---|---|---|---|---|---|---|---|---|---|---|---|---|---|
| White European | 337,692 | 62.6 (14) | 36.2% (122,106) | 21% (71,072) | 19.3% (65,134) | 12.4% (41,820) | 42.8% (144,505) | 27.5% (92,926) | 31.1% (104,934) | 22.9% (77,309) | 17.4% (58,593) | 12,227 (3.6%) | 3.6% | 1,440,897 |
| **South Asian** | **18,118** | **55.3 (13.9)** | **29.1% (5,267)** | **10.4% (1,879)** | **30.1% (5,460)** | **32.5% (5,884)** | **43.7% (7,922)** | **35.7% (6,465)** | **25.2% (4,563)** | **20.1% (3,646)** | **19.7% (3,568)** | **1,954 (10.8%)** | **10.8%** | **73,719** |
| – Indian | 9,643 | 57.6 (13.5) | 31% (2,991) | 14.5% (1,401) | 20% (1,933) | 32.6% (3,140) | 48.4% (4,671) | 36.2% (3,489) | 20% (1,928) | 20.1% (1,939) | 19.8% (1,908) | 722 (7.5%) | 7.5% | 39,304 |
| – Pakistani | 6,131 | 53.2 (13.9) | 28.6% (1,755) | 6.4% (390) | 40.3% (2,470) | 31.8% (1,947) | 37.6% (2,308) | 33.7% (2,064) | 27.9% (1,708) | 23.6% (1,447) | 20.5% (1,255) | 800 (13%) | 13% | 24,966 |
| – Bangladeshi | 2,344 | 51.4 (13.9) | 22.2% (521) | 3.8% (88) | 45.1% (1,057) | 34% (797) | 40.2% (943) | 38.9% (912) | 39.5% (927) | 11.1% (260) | 17.3% (405) | 432 (18.4%) | 18.4% | 9,448 |
| Other Asian | 4,565 | 54.7 (13.4) | 27.4% (1,251) | 14.3% (654) | 17.7% (810) | 28.9% (1,319) | 41% (1,873) | 35.1% (1,601) | 24.3% (1,111) | 18.4% (841) | 18.2% (831) | 503 (11%) | 11% | 18,080 |
| **BLACK** | **5,324** | **58.7 (14.5)** | **37.9% (2,018)** | **3.8% (202)** | **40.4% (2,153)** | **32.8% (1,746)** | **57.9% (3,082)** | **35.2% (1,874)** | **28.7% (1,530)** | **31.5% (1,679)** | **9.5% (505)** | **445 (8.4%)** | **8.4%** | **21,571** |
| – African | 2,177 | 53.3 (13.3) | 31.5% (685) | 4.4% (95) | 40% (870) | 27.5% (598) | 47.9% (1,043) | 29.8% (648) | 26.4% (575) | 30.3% (659) | 8.9% (194) | 296 (13.6%) | 13.6% | 8,264 |
| – Caribbean | 3,147 | 62.4 (14.1) | 42.4% (1,333) | 3.4% (107) | 40.8% (1,283) | 36.5% (1,148) | 64.8% (2,039) | 39% (1,226) | 30.3% (955) | 32.4% (1,020) | 9.9% (311) | 149 (4.7%) | 4.7% | 13,307 |
| Other Black | 730 | 54 (14.2) | 30.8% (225) | 7.3% (53) | 39% (285) | 24.4% (178) | 46.6% (340) | 27.4% (200) | 35.5% (259) | 29% (212) | 11.9% (87) | 77 (10.5%) | 10.5% | 2,913 |
| Chinese | 680 | 59.1 (13.5) | 29.3% (199) | 19.4% (132) | 19.1% (130) | 21.2% (144) | 43.4% (295) | 28.5% (194) | 22.9% (156) | 9.4% (64) | 9.9% (67) | 42 (6.2%) | 6.2% | 2,863 |
| Mixed | 1,725 | 55.5 (14.2) | 32.3% (558) | 17.3% (298) | 25.1% (433) | 19.8% (342) | 40% (690) | 30.1% (519) | 34.8% (600) | 23.7% (409) | 16.5% (285) | 193 (11.2%) | 11.2% | 7,158 |
| Other | 3,864 | 55.5 (13.7) | 27.4% (1,059) | 9.7% (376) | 38.2% (1,476) | 17.3% (668) | 32.1% (1,242) | 26.8% (1,037) | 32.6% (1,258) | 23.5% (909) | 17.9% (691) | 372 (9.6%) | 9.6% | 15,204 |
| Unknown/Missing | 4,297 | 67.9 (17.2) | 38.5% (1,655) | 23.4% (1,004) | 16% (687) | 10.8% (464) | 43.2% (1,855) | 17.2% (738) | 27.3% (1,174) | 13% (560) | 6.9% (298) | 188 (4.4%) | 4.4% | 13,511 |

*N = 360,994. CAD = Coronary Artery Disease; IMD = Index of Multiple Deprivation; BMI = Body Mass Index. Premature CAD = first CAD event ≤45 years. Non-premature CAD = CAD events in those aged >45 years at first event.*

Table S2: Baseline characteristics by ethnic group, non premature CAD cohort, n=360,994

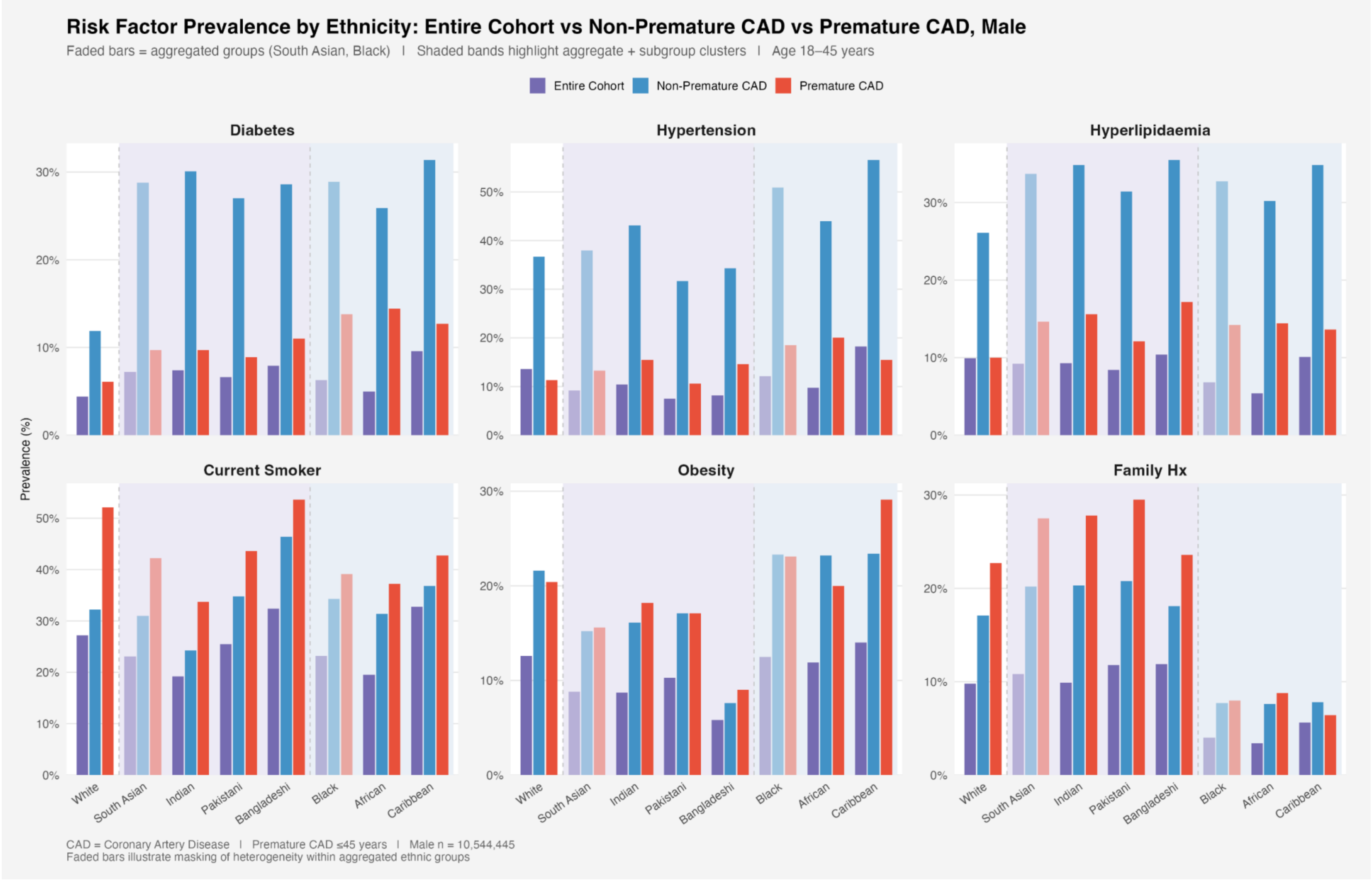

Figure S1: Cardiovascular risk factor prevalence by ethnic group, entire cohort, non premature CAD, and premature CAD patients— **male**

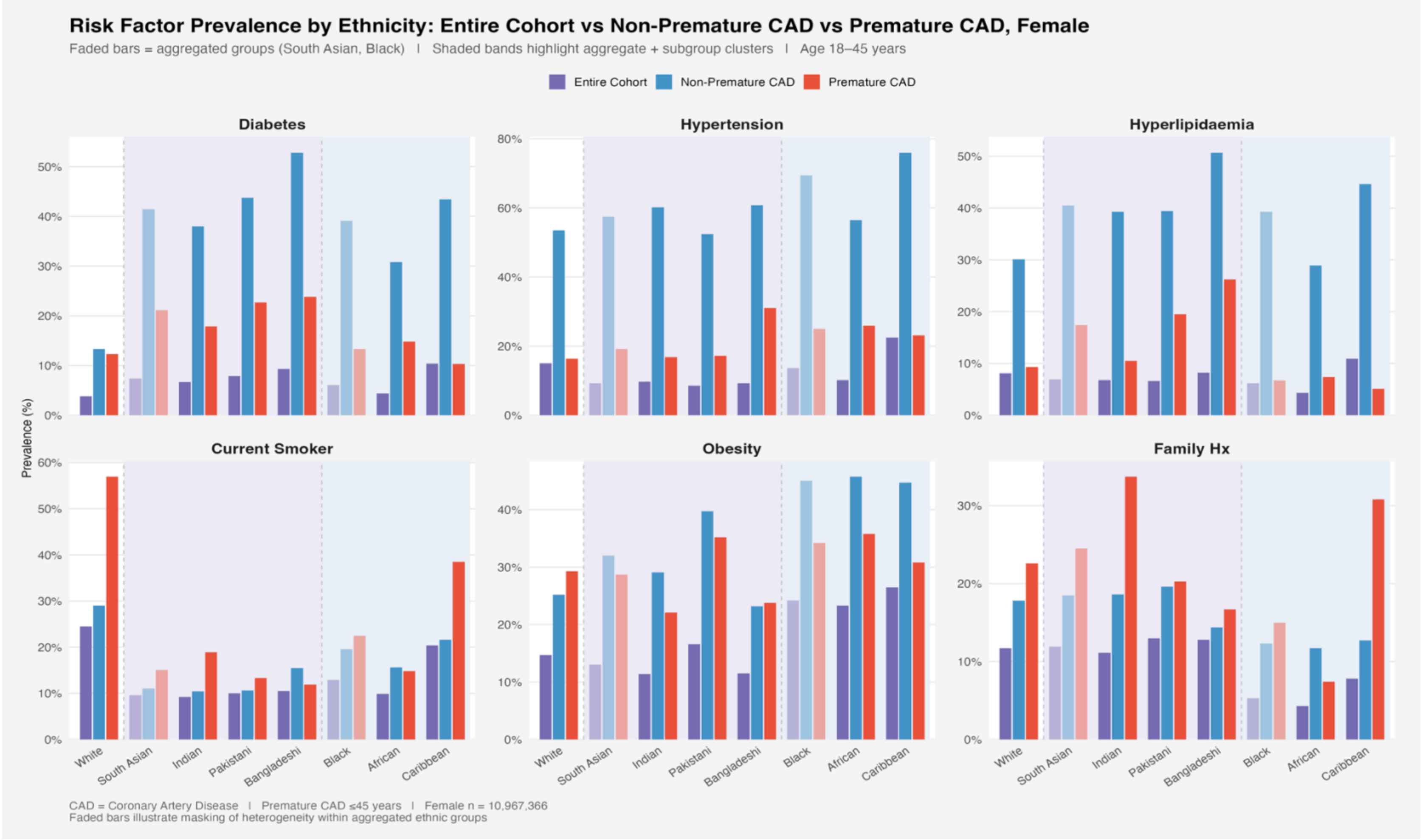

Figure S2: Cardiovascular risk factor prevalence by ethnic group, entire cohort, non premature CAD, and premature CAD patients— Female

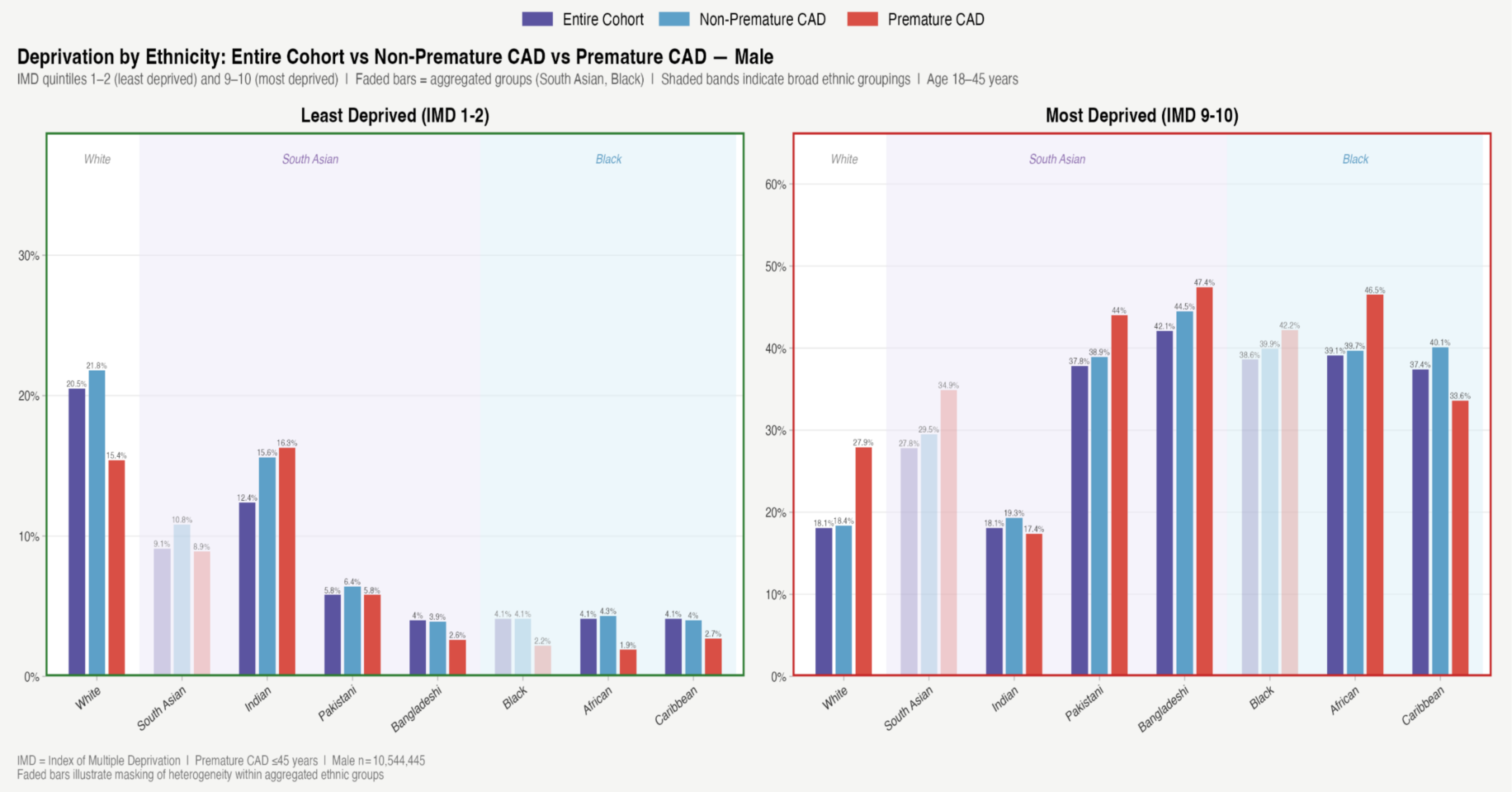

Figure S3: Deprivation by Ethnicity — Men

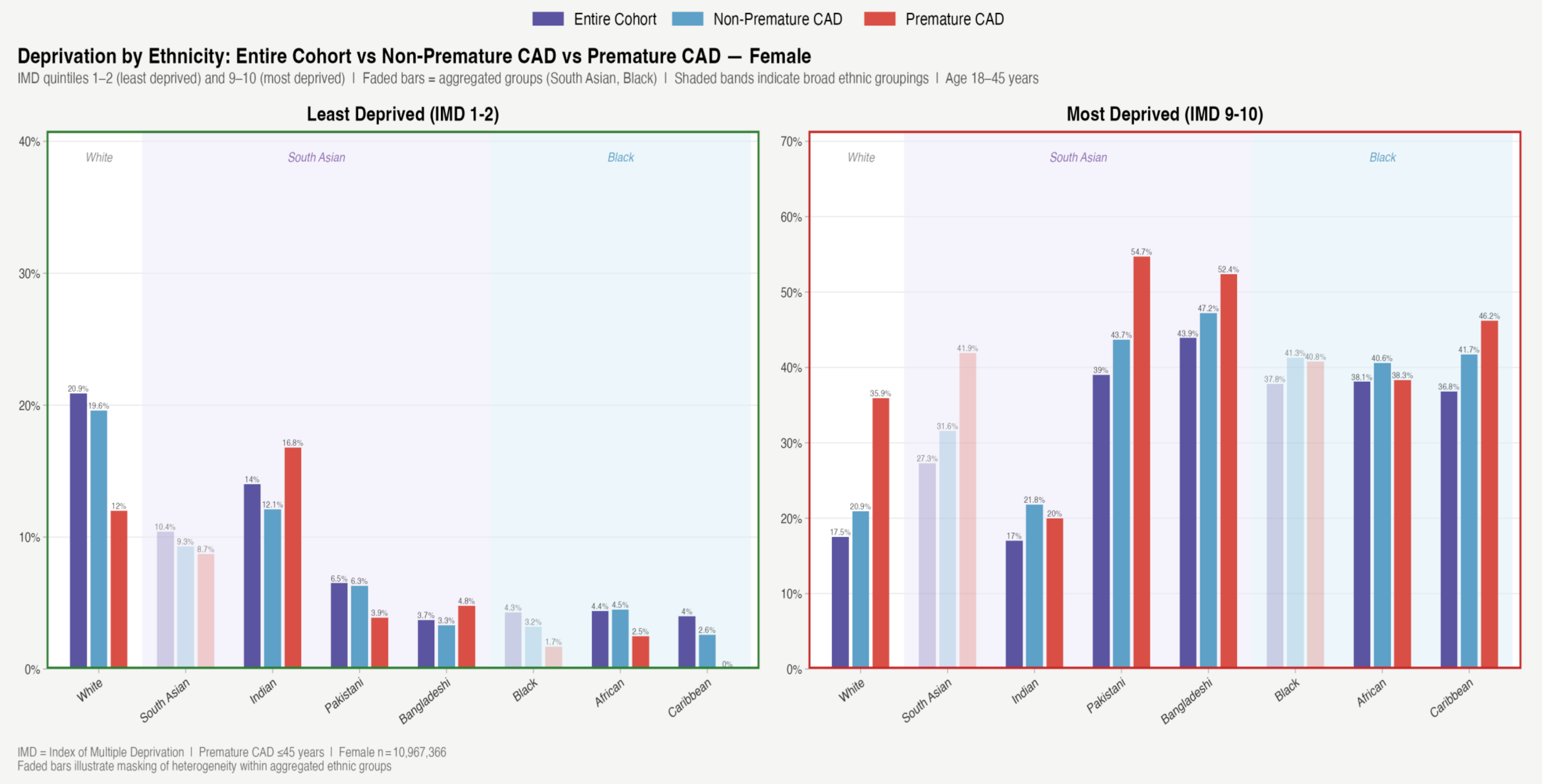

Figure S4: Deprivation by Ethnicity — Women

| Ethnic Group (18-45) | Age adjusted IRR (95% CI) | Fully adjusted IRR | Age adjusted IRR | Fully adjusted IRR | Age Adjusted IRR | Fully adjusted IRR | Age adjusted IRR | Fully Age adjusted IRR |
|---|---|---|---|---|---|---|---|---|
| | Main complete case | Main complete full case | Paired | Paired | Including unknown | Including unknown | Complete BMI | Complete BMI |
| South Asian | 1.87 (1.77, 1.99) | 1.74 (1.64, 1.84) | 1.87 (1.77, 1.99) | 1.75 (1.65, 1.86) | 1.87 (1.77, 1.99) | 1.74 (1.64, 1.85) | 1.81(1,69, 1.94) | 1.75(1.64, 1.87) |
| Indian | 1.39 (1.28, 1.52) | 1.43 (1.32, 1.56) | 1.39 (1.28, 1.50) | 1.45 (1.33, 1.57) | 1.39 (1.28, 1.52) | 1.43 (1.32, 1.56) | 1.30(1.18, 1.43) | 1.41(1.28, 1.55) |
| Pakistani | 2.16 (1.99, 2.33) | 1.90 (1.75, 2.07) | 2.16 (2.00, 2.33) | 1.91 (1.75, 2.07) | 2.16 (1.99, 2.34) | 1.90 (1.75, 2.07) | 2.15(1.95, 2.37) | 1.94(1.76, 2.14) |
| Bangladeshi | 2.79 (2.52, 3.07) | 2.20 (1.98, 2.46) | 2.79 (2.52, 3.07) | 2.21 (1.97, 2.46) | 2.79 (2.52, 3.07) | 2.20 (1.98, 2.46) | 2.79(2.48, 3.13) | 2.27(2.01, 2.57) |
| Black | 0.64 (0.57, 0.71) | 0.66 (0.59, 0.73) | 0.64 (0.57, 0.71) | 0.65 (0.59, 0.73) | 0.64 (0.57, 0.71) | 0.66 (0.59, 0.73) | 0.62(0.55, 0.70) | 0.67(0.59, 0.76) |
| African | 0.57 (0.50, 0.65) | 0.62 (0.54, 0.71) | 0.57 (0.50, 0.65) | 0.62 (0.54, 0.70) | 0.57 (0.50, 0.65) | 0.62 (0.54, 0.71) | 0.55(0.48, 0.64) | 0.63 (0.54, 0.73) |
| Caribbean | 0.82 (0.68, 0.98) | 0.75 (0.63, 0.90) | 0.82 (0.68, 0.98) | 0.74 (0.62, 0.89) | 0.82 (0.68, 0.98) | 0.75 (0.63, 0.90) | 0.86(0.70, 1.06) | 0.79(0.64, 0.97) |
| Other Black | 0.52 (0.40, 0.68) | 0.54 (0.41, 0.70) | | | 0.52 (0.40, 0.68) | 0.54 (0.42, 0.70) | 0.56(0.41, 0.76) | 0.59(0.43, 0.80) |
| Chinese | 0.18 (0.11, 0.29) | 0.28 (0.18, 0.44) | | | 0.18 (0.11, 0.29) | 0.28 (0.18, 0.44) | 0.14(0.08, 0.24) | 0.24(0.14, 0.40) |
| Mixed | 0.69 (0.58, 0.81) | 0.73(0.62, 0.86) | | | 0.69 (0.58, 0.81) | 0.73(0.62, 0.86) | 0.71(0.59, 0.85) | 0.79(0.66, 0.95) |
| Other Asian | 1.32 (1.17, 1.50) | 1.35 (1.20, 1.52) | | | 1.32 (1.17, 1.50) | 1.35 (1.21, 1.52) | 1.29(1.13, 1.48) | 1.40(1.23, 1.59) |
| Other | 0.92 (0.80, 1.06) | 0.88 (0.77, 1.01) | | | 0.92 (0.80, 1.06) | 0.88 (0.77, 1.01) | 0.84(0.73, 0.96) | 0.82( 0.72, 0.94) |
| Unknown/Missing | | | | | 0.09(0.07, 0.10) | 0.11(0.10, 0.13) | 0.09(0.07, 0.11) | 0.12(0.10, 0.15) |

Table S3. Male (18-45): Main Analysis and Sensitivity Analyses

| Ethnic Group (18-45) | Age adjusted IRR (95% CI) | Fully adjusted IRR | Age adjusted IRR (95% CI) | Fully adjusted IRR (95% CI) | Age adjusted IRR (95% CI) | Fully adjusted IRR (95% CI) | Age adjusted IRR (95% CI) | Fully Adjusted |
|---|---|---|---|---|---|---|---|---|
| | Main | Main | Paired | Paired | Including unknown ethnicity | Including unknown ethnicity | Complete BMI | Complete BMI |
| South Asian | 1.10 (0.95, 1.26) | 1.18 (1.02, 1.36) | 1.10 (0.95, 1.26) | 1.18 (1.03, 1.36) | 1.10 (0.95, 1.26) | 1.18 (1.02, 1.36) | 1.14(0.98, 1.33) | 1.23 (1.06, 1.44) |
| Indian | 0.78 (0.62, 0.98) | 1.01 (0.81, 1.27) | 0.78 (0.62, 0.98) | 1.02 (0.81, 1.28) | 0.78 (0.62, 0.98) | 1.01 (0.81, 1.27) | 0.79 (0.61-1.02) | 1.05 (0.82, 1.35) |
| Pakistani | 1.56 (1.29, 1.89) | 1.53 (1.26, 1.85) | 1.56 (1.29, 1.89) | 1.53 (1.26, 1.85) | 1.56 (1.29, 1.89) | 1.52 (1.26, 1.84) | 1.65 (1.35, 2.01) | 1.59 (1.30, 1.93) |
| Bangladeshi | 1.12 (0.83, 1.52) | 0.89 (0.66, 1.21) | 1.12 (0.83, 1.52) | 0.88 (0.66, 1.20) | 1.12 (0.83, 1.52) | 0.89 (0.66, 1.21) | 1.26( 0.91, 1.74) | 0.99 (0.71, 1.37) |
| Black | 0.74 (0.60, 0.90) | 0.67 (0.55, 0.82) | 0.74 (0.60, 0.90) | 0.68 (0.56, 0.83) | 0.74 (0.60, 0.90) | 0.67 (0.55, 0.82) | 0.78(0.63, 0.96) | 0.70(0.56, 0.86) |
| African | 0.68 (0.55, 0.84) | 0.69 (0.55, 0.85) | 0.68 (0.55, 0.84) | 0.70 (0.56, 0.87) | 0.68 (0.55, 0.84) | 0.69 (0.55, 0.85) | 0.73(0.58, 0.92) | 0.73(0.58, 0.93) |
| Caribbean | 0.90 (0.63, 1.28) | 0.64 (0.44, 0.91) | 0.90 (0.63, 1.28) | 0.64 (0.45, 0.91) | 0.90 (0.63, 1.28) | 0.64 (0.45, 0.91) | 0.91 (0.62, 1.34) | 0.63(0.43, 0.92) |
| Other Black | 0.64 (0.42, 0.96) | 0.57 (0.38, 0.86) | | | 0.64 (0.42, 0.96) | 0.57 (0.38, 0.86) | 0.65(0.40, 1.03) | 0.57 (0.35, 0.91) |
| Chinese | 0.19 (0.10, 0.36) | 0.40 (0.21, 0.73) | | | 0.19 (0.10, 0.36) | 0.40 (0.21, 0.73) | 0.17(0.08, 0.35) | 0.37(0.18, 0.76) |
| Mixed | 0.57 (0.41, 0.79) | 0.61 (0.44, 0.84) | | | 0.57 (0.41, 0.79) | 0.61 (0.44, 0.84) | 0.59(0.41, 0.86) | 0.63(0.43, 0.91) |
| Other Asian | 0.77 (0.62, 0.96) | 1.03 (0.83, 1.28) | | | 0.77 (0.62, 0.96) | 1.03 (0.83, 1.28) | 0.79(0.62, 1.01) | 1.08 (0.85, 1.37) |
| Other | 0.66 (0.46, 0.95) | 0.75 (0.53, 1.07) | | | 0.66 (0.46, 0.95) | 0.75 (0.53, 1.07) | 0.59(0.43, 0.80) | 0.67 (0.49, 0.91) |
| Unknown/Missing | | | | | 0.09 (0.06, 0.14) | 0.14(0.09, 0.22) | 0.09(0.05, 0.16) | 0.15 (0.08, 0.26) |

Table S4. Female (18-45): Main Analysis and Sensitivity Analyses

**Male participants aged 18-45**

| Ethnic group (18-45), male | Model 1 Age Adjusted | Model 2 Age adjusted + IMD | Model 3 Age adjusted + DM | Model 4 Age adjusted + Smoking | Model 5 Age adjusted + HTN | Model 6 Age adjusted + Hyperlipidaemia | Model 7 Age adjusted + family history | Model 8 Age adjusted + BMI | Fully adjusted model |
|---|---|---|---|---|---|---|---|---|---|
| South Asian | 1.87 (1.77, 1.99) | 1.73 (1.63, 1.83) | 1.80 (1.70, 1.91) | 2.01 (1.90, 2.14) | 1.85 (1.74, 1.96) | 1.71 (1.61, 1.81) | 1.77 (1.67, 1.88) | 1.90 (1.78, 2.02) | 1.75 (1.65, 1.86) |
| Indian | 1.39 (1.28, 1.50) | 1.36 (1.25, 1.46) | 1.31 (1.21, 1.43) | 1.59 (1.46, 1.74) | 1.35 (1.24, 1.47) | 1.27 (1.16, 1.38) | 1.34 (1.23, 1.47) | 1.39 (1.28, 1.52) | 1.45 (1.33, 1.57) |
| Pakistani | 2.16 (2.00, 2.33) | 1.92 (1.77, 2.08) | 2.10 (1.93, 2.28) | 2.26 (2.07, 2.45) | 2.17 (2.00, 2.36) | 2.02 (1.86, 2.19) | 2.00 (1.84, 2.18) | 2.17 (1.99, 2.36) | 1.91 (1.75, 2.07) |
| Bangladeshi | 2.79 (2.52, 3.07) | 2.38 (2.17, 2.64) | 2.65 (2.40, 2.91) | 2.66 (2.40, 2.95) | 2.75 (2.49, 3.03) | 2.41 (2.18, 2.67) | 2.59 (2.34, 2.87) | 2.99 (2.71, 3.30) | 2.21 (1.97, 2.46) |
| Black | 0.64 (0.57, 0.71) | 0.55 (0.49, 0.61) | 0.62 (0.56, 0.69) | 0.70 (0.63, 0.77) | 0.60 (0.53, 0.66) | 0.63 (0.56, 0.70) | 0.69 (0.62, 0.77) | 0.61(0.55, 0.68) | 0.65 (0.59, 0.73) |
| African | 0.57 (0.50, 0.65) | 0.49 (0.43, 0.56) | 0.56 (0.49, 0.63) | 0.65 (0.57, 0.75) | 0.53 (0.46, 0.61) | 0.56 (0.49, 0.64) | 0.63 (0.55, 0.72) | 0.55 (0.48, 0.63) | 0.62 (0.54, 0.70) |
| Caribbean | 0.82 (0.68, 0.98) | 0.71 (0.59, 0.85) | 0.80 (0.67, 0.96) | 0.80 (0.67, 0.95) | 0.78 (0.65, 0.93) | 0.83 (0.69, 0.99) | 0.87 (0.73, 1.04) | 0.80 (0.67, 0.95) | 0.74 (0.62, 0.89) |

**Table S5:** Paired Analysis 18-45 Male using Poisson regression models.

**Female participants aged 18-45**

| Ethnic group (18-45), female | Model 1 Age adjusted | Model 2 Age adjusted + IMD | Model 3 Age adjusted + DM | Model 4 Age adjusted + Smoking | Model 5 Age adjusted + HTN | Model 6 Age adjusted + Hyperlipidaemia | Model 7 Age adjusted + family history | Model 8 Age adjusted + BMI | Fully adjusted model |
|---|---|---|---|---|---|---|---|---|---|
| South Asian | 1.10 (0.95, 1.26) | 0.94 (0.82, 1.08) | 0.99 (0.86, 1.14) | 1.54 (1.34, 1.77) | 1.11 (0.97, 1.28) | 1.03 (0.90, 1.19) | 1.07 (0.93, 1.23) | 1.13 (0.98, 1.31) | 1.18 (1.03, 1.36) |
| Indian | 0.78 (0.62, 0.98) | 0.76 (0.61, 0.96) | 0.72 (0.57, 0.91) | 1.06 (0.84, 1.34) | 0.80(0.64, 1.01) | 0.75 (0.60, 0.95) | 0.77 (0.61, 0.97) | 0.83 (0.66, 1.04) | 1.02 (0.81, 1.28) |
| Pakistani | 1.56 (1.29, 1.89) | 1.21 (0.99, 1.46) | 1.40 (1.15, 1.69) | 2.17 (1.79, 2.63) | 1.60 (1.32, 1.93) | 1.49 (1.23, 1.81) | 1.50 (1.24, 1.81) | 1.54 (1.27, 1.86) | 1.53 (1.26, 1.85) |
| Bangladeshi | 1.12 (0.83, 1.52) | 0.82 (0.60, 1.11) | 0.95 (0.70, 1.27) | 1.56 (1.16, 2.10) | 1.07 (0.80, 1.45) | 0.95 (0.71, 1.29) | 1.08 (0.80, 1.46) | 1.16 (0.86, 1.57) | 0.88 (0.66, 1.20) |
| Black | 0.74 (0.60, 0.90) | 0.57 (0.46, 0.69) | 0.71 (0.58, 0.87) | 0.96 (0.79, 1.18) | 0.65 (0.53, 0.79) | 0.72 (0.59, 0.87) | 0.79 (0.65, 0.96) | 0.63 (0.54, 0.81) | 0.68 (0.56, 0.83) |
| African | 0.68 (0.55, 0.84) | 0.52 (0.42, 0.65) | 0.66 (0.53, 0.82) | 0.96 (0.77, 1.20) | 0.61 (0.49, 0.75) | 0.66 (0.53, 0.83) | 0.74 (0.59, 0.92) | 0.58 (0.46, 0.72) | 0.70 (0.56, 0.87) |
| Caribbean | 0.90 (0.63, 1.28) | 0.69 (0.48, 0.98) | 0.85 (0.59, 1.21) | 0.97 (0.68, 1.39) | 0.76 (0.53, 1.08) | 0.86 (0.60, 1.23) | 0.94 (0.65, 1.34) | 0.77 (0.54, 1.10) | 0.64 (0.45, 0.91) |

**Table S6:** Paired Analysis 18-45 Female using Poisson regression models.

| Ethnic group (18-45), male | Model 1 Age standardised | Model 2 + IMD | Model 3 + DM | Model 4 + Smoking | Model 5 + HTN | Model 6 + Hyperlipidaemia | Model 7 + family hx | Model 8 + BMI |
|---|---|---|---|---|---|---|---|---|
| South Asian | 1.87 (1.77, 1.99) | 1.74 (1.64, 1.84) | 1.67 (1.58, 1.77) | 1.84 (1.73, 1.95) | 1.83 (1.72, 1.93) | 1.75 (1.65, 1.85) | 1.68 (1.59, 1.78) | 1.74 (1.64, 1.84) |
| Indian | 1.39 (1.28, 1.52) | 1.36 (1.25, 1.49) | 1.31 (1.19, 1.43) | 1.50 (1.38, 1.64) | 1.47 (1.35, 1.61) | 1.42 (1.30, 1.55) | 1.39 (1.28, 1.52) | 1.43 (1.32, 1.56) |
| Pakistani | 2.16 (1.99, 2.33) | 1.93 (1.78, 2.09) | 1.87 (1.72, 2.03) | 2.02 (1.86, 2.20) | 2.05 (1.88, 2.23) | 1.98 (1.82, 2.15) | 1.87 (1.72, 2.04) | 1.90 (1.75, 2.07) |
| Bangladeshi | 2.79 (2.52, 3.07) | 2.41 (2.18, 2.66) | 2.29 (2.08, 2.53) | 2.31 (2.08, 2.55) | 2.30 (2.09, 2.55) | 2.12 (1.91, 2.35) | 2.02 (1.81, 2.25) | 2.20 (1.98, 2.46) |
| Black | 0.64 (0.57, 0.71) | 0.56 (0.50, 0.62) | 0.55 (0.49, 0.61) | 0.62 (0.56, 0.69) | 0.60 (0.54, 0.67) | 0.61 (0.54, 0.68) | 0.66 (0.59, 0.73) | 0.66 (0.59, 0.73) |
| African | 0.57 (0.50, 0.65) | 0.50 (0.44, 0.58) | 0.49 (0.43, 0.56) | 0.58 (0.51, 0.67) | 0.56 (0.49, 0.64) | 0.56 (0.49, 0.64) | 0.62 (0.54, 0.70) | 0.62 (0.54, 0.71) |
| Caribbean | 0.82 (0.68, 0.98) | 0.73 (0.61, 0.87) | 0.72 (0.60, 0.86) | 0.73 (0.61, 0.87) | 0.70 (0.58, 0.84) | 0.72 (0.60, 0.86) | 0.76 (0.64, 0.91) | 0.75 (0.63, 0.90) |
| Other Black | 0.52 (0.40, 0.68) | 0.47 (0.36, 0.61) | 0.46 (0.36, 0.61) | 0.50 (0.39, 0.65) | 0.49 (0.38, 0.64) | 0.50 (0.39, 0.65) | 0.54 (0.41, 0.70) | 0.54 (0.41, 0.70) |
| Chinese | 0.18 (0.11, 0.29) | 0.18 (0.11, 0.28) | 0.18(0.12, 0.29) | 0.22 (0.14, 0.34) | 0.23 (0.15, 0.36) | 0.23 (0.15, 0.36) | 0.25 (0.16, 0.39) | 0.28 (0.18, 0.44) |
| Mixed | 0.69 (0.58, 0.81) | 0.65 (0.55, 0.76) | 0.65(0.56, 0.77) | 0.67 (0.57, 0.79) | 0.68 (0.58, 0.80) | 0.69 (0.59, 0.81) | 0.72 (0.61, 0.84) | 0.73(0.62, 0.86) |
| Other Asian | 1.32 (1.17, 1.50) | 1.28 (1.13, 1.45) | 1.26 (1.11, 1.42) | 1.37 (1.201, 1.54) | 1.36 (1.21, 1.53) | 1.30 (1.15, 1.46) | 1.31 (1.17, 1.47) | 1.35 (1.20, 1.52) |
| Other | 0.92 (0.80, 1.06) | 0.85 (0.74, 0.97) | 0.86 (0.75, 0.98) | 0.87(0.76, 0.99) | 0.88 (0.77, 1.01) | 0.87 (0.76, 0.99) | 0.90 (0.78, 1.03) | 0.88 (0.77, 1.01) |

Table S7: Sequential models 18-45 Male

| Ethnic group (18-45), female | Model 1 Age standardised | Model 2 + IMD | Model 3 + DM | Model 4 + Smoking | Model 5 + HTN | Model 6 + Hyperlipidaemia | Model 7 + family hx | Model 8 BMI |
|---|---|---|---|---|---|---|---|---|
| South Asian | 1.10 (0.95, 1.26) | 0.94 (0.82, 1.09) | 0.86 (0.74, 0.99) | 1.20 (1.05, 1.39) | 1.21 (1.05, 1.39) | 1.17 (1.02, 1.35) | 1.14 (0.99, 1.31) | 1.18 (1.02, 1.36) |
| Indian | 0.78 (0.62, 0.98) | 0.76 (0.61, 0.96) | 0.71 (0.57, 0.90) | 0.98 (0.78, 1.23) | 1.00 (0.79, 1.25) | 0.98 (0.78, 1.24) | 0.97 (0.77, 1.21) | 1.01 (0.81, 1.27) |
| Pakistani | 1.56 (1.29, 1.89) | 1.22 (1.01, 1.48) | 1.10 (0.92, 1.34) | 1.57 (1.30, 1.90) | 1.61 (1.32, 1.94) | 1.57 (1.30, 1.90) | 1.52 (1.25, 1.83) | 1.53 (1.26, 1.85) |
| Bangladeshi | 1.12 (0.83, 1.52) | 0.83 (0.61, 1.12) | 0.71 (0.52, 0.95) | 1.01 (0.75, 1.36) | 0.96 (0.71, 1.29) | 0.87 (0.64, 1.17) | 0.84 (0.62, 1.13) | 0.89 (0.66, 1.21) |
| Black | 0.74 (0.60, 0.90) | 0.57 (0.47, 0.69) | 0.55 (0.46, 0.67) | 0.75 (0.62, 0.91) | 0.67 (0.55, 0.82) | 0.68 (0.56, 0.82) | 0.72 (0.59, 0.87) | 0.67 (0.55, 0.82) |
| African | 0.68 (0.55, 0.84) | 0.53 (0.42, 0.65) | 0.52 (0.41, 0.64) | 0.74 (0.60, 0.93) | 0.68 (0.55, 0.85) | 0.69 (0.55, 0.86) | 0.74 (0.59, 0.92) | 0.69 (0.55, 0.85) |
| Caribbean | 0.90 (0.63, 1.28) | 0.70 (0.49, 0.99) | 0.66 (0.47, 0.95) | 0.76 (0.53, 1.08) | 0.65 (0.45, 0.93) | 0.66 (0.46, 0.94) | 0.68 (0.48, 0.97) | 0.64 (0.44, 0.91) |
| Other Black | 0.64 (0.42, 0.96) | 0.50 (0.34, 0.75) | 0.50 (0.33, 0.75) | 0.60 (0.40, 0.90) | 0.56 (0.37, 0.84) | 0.57 (0.38, 0.86) | 0.59 (0.39, 0.89) | 0.57 (0.38, 0.86) |
| Chinese | 0.19 (0.10, 0.36) | 0.19 (0.10, 0.36) | 0.21 (0.11, 0.39) | 0.29 (0.16, 0.54) | 0.31 (0.17, 0.58) | 0.31 (0.17, 0.58) | 0.33 (0.18, 0.61) | 0.40 (0.21, 0.73) |
| Mixed | 0.57 (0.41, 0.79) | 0.52 (0.37, 0.72) | 0.52 (0.37, 0.72) | 0.56 (0.41, 0.78) | 0.58 (0.42, 0.80) | 0.58 (0.42, 0.81) | 0.60 (0.43, 0.83) | 0.61 (0.44, 0.84) |
| Other Asian | 0.77 (0.62, 0.96) | 0.75 (0.60, 0.93) | 0.72 (0.58, 0.90) | 0.98 (0.79, 1.23) | 1.00 (0.80, 1.24) | 0.97 (0.78, 1.21) | 0.97 (0.78, 1.21) | 1.03 (0.83, 1.28) |
| Other | 0.66 (0.46, 0.95) | 0.57 (0.41, 0.81) | 0.58 (0.41, 0.82) | 0.70 (0.49, 0.99) | 0.73 (0.51, 1.04) | 0.73 (0.51, 1.03) | 0.74 (0.52, 1.05) | 0.75 (0.53, 1.07) |

Table S8: Sequential models 18-45 Females

| **Ethnic group (18-45), male** | Model 1 Age adjusted | Model 2 Age adjusted + IMD | Model 3 Age adjusted + DM | Model 4 Age adjusted + Smoking | Model 5 Age adjusted + HTN | Model 6 Age adjusted + Hyperlipidaemia | Model 7 Age adjusted + family history | Model 8 Age adjusted + BMI |
|---|---|---|---|---|---|---|---|---|
| South Asian | 1.87 (1.77, 1.99) | 1.74 (1.64, 1.84) | 1.80 (1.70, 1.91) | 2.01 (1.89, 2.13) | 1.85 (1.74, 1.96) | 1.71 (1.61, 1.81) | 1.77 (1.67, 1.88) | 1.90 (1.78, 2.01) |
| Indian | 1.39 (1.28, 1.52) | 1.36 (1.25, 1.49) | 1.33 (1.22, 1.45) | 1.58 (1.45, 1.73) | 1.35 (1.24, 1.47) | 1.27 (1.16, 1.38) | 1.34 (1.23, 1.47) | 1.39 (1.28, 1.52) |
| Pakistani | 2.16 (1.99, 2.33) | 1.93 (1.78, 2.09) | 2.10 (1.93, 2.28) | 2.25 (2.07, 2.45) | 2.17 (2.00, 2.36) | 2.02 (1.86, 2.19) | 2.00 (1.84, 2.18) | 2.17 (1.99, 2.36) |
| Bangladeshi | 2.79 (2.52, 3.07) | 2.41 (2.18, 2.66) | 2.64 (2.39, 2.91) | 2.66 (2.40, 2.95) | 2.75 (2.49, 3.03) | 2.40 (2.17, 2.66) | 2.59 (2.33, 2.87) | 2.99 (2.71, 3.30) |
| Black | 0.64 (0.57, 0.71) | 0.56 (0.50, 0.62) | 0.62 (0.56, 0.69) | 0.69 (0.62, 0.77) | 0.59 (0.53, 0.66) | 0.63 (0.56, 0.70) | 0.69 (0.62, 0.77) | 0.61(0.55, 0.68) |
| African | 0.57 (0.50, 0.65) | 0.50 (0.44, 0.58) | 0.56 (0.49, 0.63) | 0.65 (0.57, 0.74) | 0.53 (0.46, 0.61) | 0.56 (0.49, 0.64) | 0.63 (0.55, 0.72) | 0.55 (0.48, 0.62) |
| Caribbean | 0.82 (0.68, 0.98) | 0.73 (0.61, 0.87) | 0.80 (0.67, 0.96) | 0.80 (0.67, 0.95) | 0.78 (0.65, 0.93) | 0.83 (0.69, 0.99) | 0.87 (0.73, 1.04) | 0.80 (0.67, 0.96) |
| Other Black | 0.52 (0.40, 0.68) | 0.47 (0.36, 0.61) | 0.52 (0.40, 0.68) | 0.54 (0.42, 0.71) | 0.52 (0.40, 0.67) | 0.53 (0.41, 0.69) | 0.56 (0.43, 0.74) | 0.52 (0.40, 0.68) |
| Chinese | 0.18 (0.11, 0.29) | 0.18 (0.11, 0.28) | 0.19(0.12, 0.30) | 0.21(0.14, 0.33) | 0.19 (0.12, 0.31) | 0.19 (0.12, 0.30) | 0.20 (0.13, 0.32) | 0.20 (0.13, 0.32) |
| Mixed | 0.69 (0.58, 0.81) | 0.65 (0.55, 0.76) | 0.69(0.59, 0.81) | 0.69 (0.58, 0.81) | 0.70 (0.60, 0.82) | 0.69 (0.59, 0.82) | 0.72 (0.61, 0.84) | 0.70(0.59, 0.82) |
| Other Asian | 1.32 (1.17, 1.50) | 1.28 (1.13, 1.45) | 1.30 (1.15, 1.46) | 1.41 (1.25, 1.59) | 1.31 (1.17, 1.48) | 1.22 (1.09, 1.38) | 1.33 (1.18, 1.49) | 1.33 (1.18, 1.50) |
| Other | 0.92 (0.80, 1.06) | 0.85 (0.74, 0.97) | 0.93 (0.81, 1.06) | 0.90(0.79, 1.03) | 0.94 (0.82, 1.08) | 0.90 (0.79, 1.04) | 0.95 (0.83, 1.09) | 0.89 (0.77, 1.02) |

Table S9: Single-covariate models, male 18–45

| Ethnic group (18-45), female | Model 1 Age adjusted | Model 2 Age adjusted + IMD | Model 3 Age adjusted + DM | Model 4 Age adjusted + Smoking | Model 5 Age adjusted + HTN | Model 6 Age adjusted + Hyperlipidaemia | Model 7 Age adjusted + family history | Model 8 Age adjusted + BMI |
|---|---|---|---|---|---|---|---|---|
| South Asian | 1.10 (0.95, 1.26) | 0.94 (0.82, 1.09) | 0.99 (0.86, 1.14) | 1.52 (1.32, 1.75) | 1.11 (0.97, 1.28) | 1.04 (0.90, 1.20) | 1.07 (0.93, 1.23) | 1.14 (0.99, 1.31) |
| Indian | 0.78 (0.62, 0.98) | 0.76 (0.61, 0.96) | 0.72 (0.57, 0.91) | 1.08 (0.86, 1.36) | 0.80(0.64, 1.01) | 0.75 (0.60, 0.95) | 0.77 (0.61, 0.97) | 0.83 (0.66, 1.04) |
| Pakistani | 1.56 (1.29, 1.89) | 1.22 (1.01, 1.48) | 1.40 (1.15, 1.69) | 2.15 (1.77, 2.60) | 1.60 (1.32, 1.93) | 1.50 (1.24, 1.81) | 1.50 (1.23, 1.81) | 1.54 (1.27, 1.87) |
| Bangladeshi | 1.12 (0.83, 1.52) | 0.83 (0.61, 1.12) | 0.95 (0.70, 1.27) | 1.54 (1.15, 2.08) | 1.07 (0.80, 1.45) | 0.96 (0.71, 1.30) | 1.08 (0.80, 1.46) | 1.16 (0.86, 1.57) |
| Black | 0.74 (0.60, 0.90) | 0.57 (0.47, 0.69) | 0.71 (0.58, 0.87) | 0.95 (0.78, 1.16) | 0.64 (0.53, 0.79) | 0.72 (0.59, 0.87) | 0.79 (0.65, 0.97) | 0.63 (0.51, 0.76) |
| African | 0.68 (0.55, 0.84) | 0.53 (0.42, 0.65) | 0.66 (0.53, 0.82) | 0.94 (0.76, 1.17) | 0.60 (0.48, 0.75) | 0.66 (0.53, 0.83) | 0.74 (0.59, 0.92) | 0.57 (0.46, 0.71) |
| Caribbean | 0.90 (0.63, 1.28) | 0.70 (0.49, 0.99) | 0.85 (0.59, 1.21) | 0.97 (0.68, 1.38) | 0.75 (0.53, 1.07) | 0.86 (0.60, 1.23) | 0.94 (0.65, 1.34) | 0.77 (0.54, 1.10) |
| Other Black | 0.64 (0.42, 0.96) | 0.50 (0.34, 0.75) | 0.63 (0.42, 0.95) | 0.72 (0.48, 1.099) | 0.59 (0.39, 0.89) | 0.64 (0.42, 0.96) | 0.68 (0.45, 1.02) | 0.58 (0.39, 0.87) |
| Chinese | 0.19 (0.10, 0.36) | 0.19 (0.10, 0.36) | 0.21 (0.11, 0.39) | 0.27 (0.15, 0.51) | 0.22 (0.12, 0.41) | 0.20 (0.11, 0.38) | 0.21 (0.11, 0.40) | 0.27 (0.14, 0.49) |
| Mixed | 0.57 (0.41, 0.79) | 0.52 (0.37, 0.72) | 0.57 (0.41, 0.79) | 0.61 (0.44, 0.84) | 0.59 (0.43, 0.82) | 0.58 (0.42, 0.80) | 0.59 (0.43, 0.82) | 0.58 (0.42, 0.81) |
| Other Asian | 0.77 (0.62, 0.96) | 0.75 (0.60, 0.93) | 0.75(0.60, 0.93) | 1.05 (0.84, 1.31) | 0.80 (0.64, 1.00) | 0.75 (0.60, 0.94) | 0.78 (0.63, 0.98) | 0.85 (0.68, 1.06) |
| Other | 0.66 (0.46, 0.95) | 0.57 (0.41, 0.81) | 0.67 (0.46, 0.96) | 0.77 (0.54, 1.10) | 0.71 (0.49, 1.02) | 0.66 (0.46, 0.95) | 0.68 (0.47, 0.97) | 0.69 (0.48, 0.99) |

Table S10: Single-covariate models, female 18–45

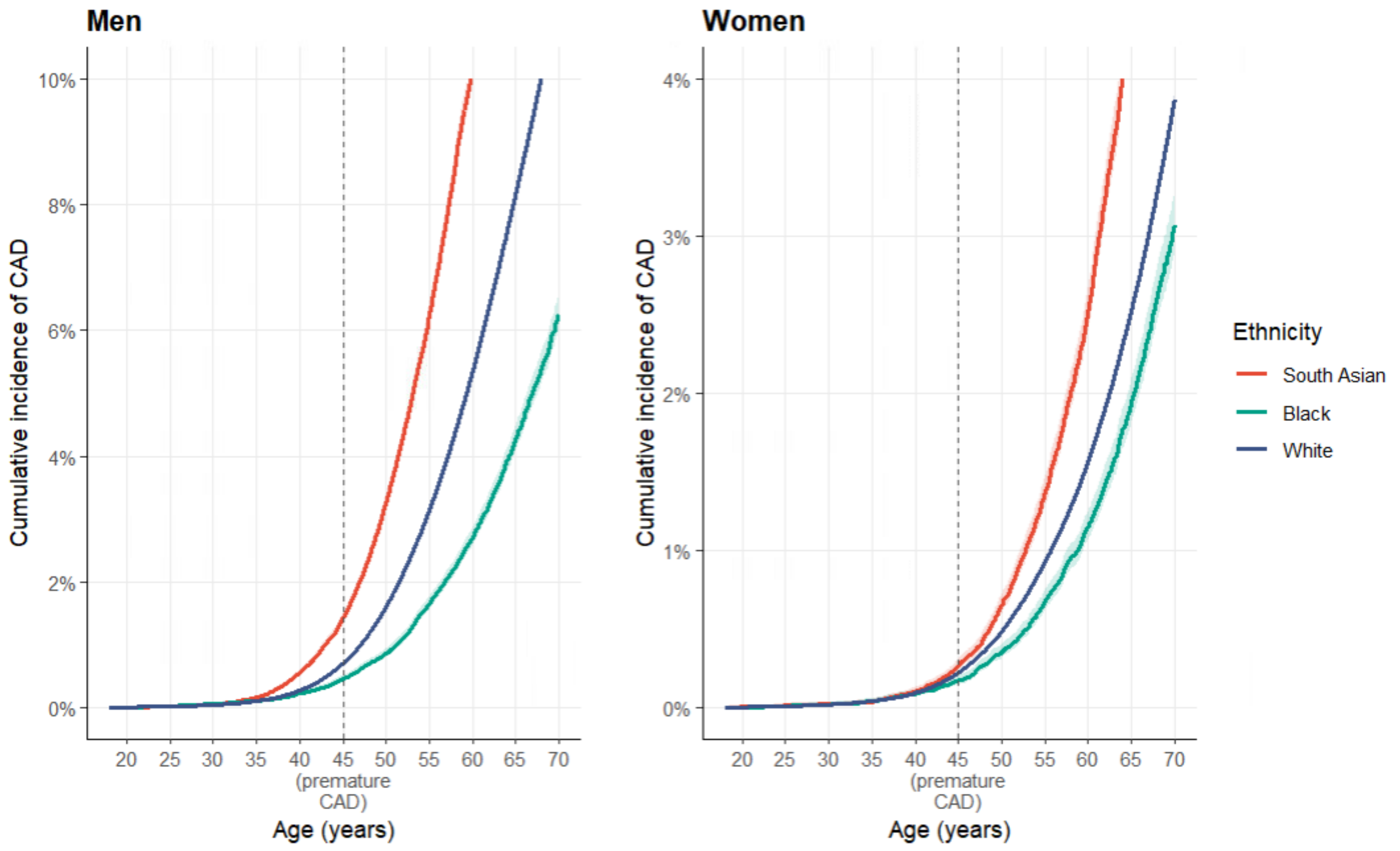

Figure S5: Cumulative incidence of CAD aggregated ethnic group.

| Ethnic Group | Sex | Census Pop (18–45) | White reference events†[1] | Age-std IRR | Excess/Deficit (age-std) | Fully adj IRR | Excess/Deficit (fully adj) |
|---|---|---|---|---|---|---|---|
| | | | | **South Asian** | | | |
| Indian | Male | 431,500 | 712 | 1.39 (1.28, 1.50) | +278[+199, +356] | 1.45 (1.33, 1.57) | +320 [+235, +406] |
| Pakistani | Male | 354,737 | 586 | 2.16 (2.00, 2.33) | +680 [+586, +779] | 1.91 (1.75, 2.07) | +533 [+440, +627] |
| Bangladeshi | Male | 146,249 | 241 | 2.79 (2.52, 3.07) | +431 [+366, +499] | 2.21 (1.97, 2.46) | +292 [+234, +352] |
| Pakistani | Female | 364,734 | 185 | 1.56 (1.29, 1.89) | +104 [+54, +165] | 1.53 (1.26, 1.85) | +98 [+48, +157] |
| Indian | Female | 431,461 | 219 | 0.78 (0.62, 0.98) | −48 [−83, −4] | 1.02 (0.81, 1.28) | +4 [−42, +61] |
| Bangladeshi | Female | 159,241 | 81 | 1.12 (0.83, 1.52) | +10 [−14, +42] | 0.88 (0.66, 1.20) | −10 [−28, +16] |
| **Total South Asian excess** | | | **NA** | | **+1,454** | | +1,238 |
| | | | | **Black** | | | |
| African | Male | 311,818 | 515 | 0.57 (0.50, 0.65) | −221 [−258, −180] | 0.62 (0.54, 0.70 | −196 [−237, −155] |
| Caribbean | Male | 105,723 | 175 | 0.82 (0.68, 0.98) | −32 [−56, −4] | 0.74 (0.62, 0.89) | −46 [−66, −19] |
| African | Female | 368,308 | 187 | 0.68 (0.55, 0.84) | −60 [−84, −30] | 0.70 (0.56, 0.87) | −56 [−82, −24] |
| Caribbean | Female | 121,749 | 62 | 0.90 (0.63, 1.28) | −6 [−23, +17] | 0.64 (0.45, 0.91) | −22 [−34, −6] |
| Total Black | | | | | -319 | | -320 |

[1] †White European crude incidence rate × Census population × 5 years (0.330/1,000 PY men; 0.102/1,000 PY women). Age-std IRR = Model 1; Fully adj IRR = Model 9.

Table S11: Expected events under White European rates, computed as the White European crude incidence rate (0.330 per 1,000 person-years in men, 0.102 in women) applied to the 2021 Census population aged 18–45 over a standardised 5-year horizon (mean observed cohort follow-up 5.46 years). Figures are an illustrative projection of population burden, not a forecast

| Male | Age band | | |
|---|---|---|---|
| Ethnic group- | 18-26 | 27-35 | 36-45 |
| **Age adjusted** | | | |
| South Asian | 1.96(1.66, 2.32) | 2.04(1.83, 2.21) | 2.00(1.85, 2.16) |
| Indian | 1.27 (0.96, 1.67) | 1.41(1.23, 1.63) | 1.40 (1.26, 1.56) |
| Pakistani | 2.57(2.07, 3.18) | 2.57 (2.26, 2.92) | 2.43(2.14, 2.75) |
| Bangladeshi | 2.52 (1.84, 3.44) | 2.95(2.52, 3.44) | 3.40(2.96, 3.89) |
| Black | 0.80(0.59, 1.08) | 0.55(0.45, 0.68) | 0.56 (0.48, 0.65) |
| African | 0.67(0.45, 0.99) | 0.48 (0.37, 0.63) | 0.51 (0.43, 0.61) |
| Caribbean | 1.11(0.69, 1.80) | 0.80 (0.58, 1.10) | 0.69(0.54, 0.89) |
| **Fully adjusted** | | | |
| South Asian | 1.79(1.51, 2,12) | 1.94(1.78, 2.13) | 1.79(1.66, 1.94) |
| Indian | 1.30(0.98, 1.71) | 1.57(1.37, 1.79) | 1.45(1.30, 1.62) |
| Pakistani | 2.15(1.72, 2.67) | 2.22(1.95, 2.51) | 1.97(1.73, 2.23) |
| Bangladeshi | 2.07(1.51, 2.84) | 2.41(2.05, 2.82) | 2.38(2.06, 2.76) |
| Black | 0.79 (0.59, 1.07) | 0.60(0.49, 0.74) | 0.56(0.48, 0.65) |
| African | 0.69(0.46, 1.03) | 0.56(0.43, 0.72) | 0.55(0.46, 0.65) |
| Caribbean | 1.03(0.64, 1.67) | 0.73(0.53, 1.01) | 0.59(0.46, 0.77) |

Table S12: IRR Male by 18-26, 27-35 and 35-45 age band

| Female | Age-band | | |
|---|---|---|---|
| Ethnic group | 18-26 | 27-35 | 36-45 |
| **Age adjusted** | | | |
| South Asian | 1.08(0.74, 1.57) | 1.19 (0.98, 1.46) | 1.17(0.95, 1.44) |
| Indian | 0.84(0.41, 1.70) | 0.78(0.59, 1.05) | 0.72(0.52, 0.99) |
| Pakistani | 1.65(1.09, 2.51) | 1.71(1.27, 2.31) | 1.96(1.47, 2.61) |
| Bangladeshi | 0.44(0.14, 1.36) | 1.71(1.13, 2.60) | 1.43 (0.82, 2.51) |
| Black | 0.70(0.42, 1.16) | 0.76(0.57, 1.03) | 0.61(0.47, 0.80) |
| African | 0.49(0.25, 0.99) | 0.74(0.55, 1.02) | 0.57(0.40, 0.79) |
| Caribbean | 1.21(0.58, 2.56) | 0.83(0.41, 1.69) | 0.73(0.46, 1.15) |
| **Fully adjusted** | | | |
| South Asian | 1.19(0.81, 1.75) | 1.26(1.04, 1.53) | 1.11(0.90, 1.38) |
| Indian | 1.05(0.52, 2.13) | 1.06(0.79, 1.41) | 0.88(0.64, 1.22) |
| Pakistani | 1.69(1.11, 2.57) | 1.52(1.13, 2.04) | 1.62(1.21, 2.16) |
| Bangladeshi | 0.44(0.14, 1.37) | 1.25(0.83, 1.89) | 0.78(0.44, 1.38) |
| Black | 0.75(0.45, 1.24) | 0.69(0.52, 0.93) | 0.50(0.38, 0.66) |
| African | 0.56(0.27, 1.13) | 0.74(0.54, 1.01) | 0.52(0.37, 0.74) |
| Caribbean | 1.12(0.53, 2.36) | 0.59(0.29, 1.19) | 0.47(0.30, 0.75) |

**Table S13:** IRR Female by 18-26, 27-35 and 35-45 age band

### Male participants aged 18-45

| Ethnic group (18-45), male | Model 1 Age Adjusted | Fully adjusted model |
|---|---|---|
| South Asian | 1.83 (1.73, 1.94) | 1.71 (1.62, 1.81) |
| Indian | 1.39 (1.27, 1.51) | 1.44 (1.33, 1.57) |
| Pakistani | 2.10 (1.94, 2.28) | 1.85 (1.70, 2.01) |
| Bangladeshi | 2.67 (2.42, 2.95) | 2.10 (1.89, 2.35) |
| Black | 0.65 (0.58, 0.72) | 0.66 (0.59, 0.74) |
| African | 0.58 (0.50, 0.65) | 0.63 (0.55, 0.72) |
| Caribbean | 0.82 (0.69, 0.98) | 0.74 (0.62, 0.88) |

**Table S14:** Incidence rate ratios for premature coronary artery disease by ethnic subgroup male, including fatal cases identified from death registration records (sensitivity analysis). Fatal CAD cases were identified from ONS death registration records (underlying cause ICD-10 [I21, I22, I23, I24.1, I25.2, I25.6]) in individuals with no corresponding CAD code in primary care or hospital records. Only deaths occurring within the observation window were included, that is, on or before the administrative censoring date (the earliest of deregistration, last data collection, and study end); deaths occurring after a person had left CPRD follow-up were excluded, as person-time was no longer observed. This added 835 fatal incident events, increasing the total from 16,001 to 16,836. Incidence rate ratios were re-estimated including these events using the same models as the primary analysis

### Female participants aged 18-45

| Ethnic group (18-45), female | Model 1 Age adjusted | Fully adjusted model |
|---|---|---|
| South Asian | 1.10 (0.95, 1.26) | 1.15 (1.00, 1.33) |
| Indian | 0.78 (0.62, 0.98) | 1.00 (0.80, 1.25) |
| Pakistani | 1.56 (1.29, 1.89) | 1.50 (1.24, 1.80) |
| Bangladeshi | 1.12 (0.83, 1.52) | 0.85 (0.63, 1.16) |
| Black | 0.76 (0.62, 0.92) | 0.70 (0.57, 0.85) |
| African | 0.68 (0.55, 0.84) | 0.70 (0.56, 0.87) |
| Caribbean | 0.97 (0.70, 1.35) | 0.69 (0.50, 0.96) |

**Table S15:** Incidence rate ratios for premature coronary artery disease by ethnic subgroup female, including fatal cases identified from death registration records (sensitivity analysis). Fatal CAD cases were identified from ONS death registration records (underlying cause ICD-10 [I21, I22, I23, I24.1, I25.2, I25.6]) in individuals with no corresponding CAD code in primary care or hospital records. Only deaths occurring within the observation window were included, that is, on or before the administrative censoring date (the earliest of deregistration, last data collection, and study end); deaths occurring after a person had left CPRD follow-up were excluded, as person-time was no longer observed. This added 835 fatal incident events, increasing the total from 16,001 to 16,836. Incidence rate ratios were re-estimated including these events using the same models as the primary analysis

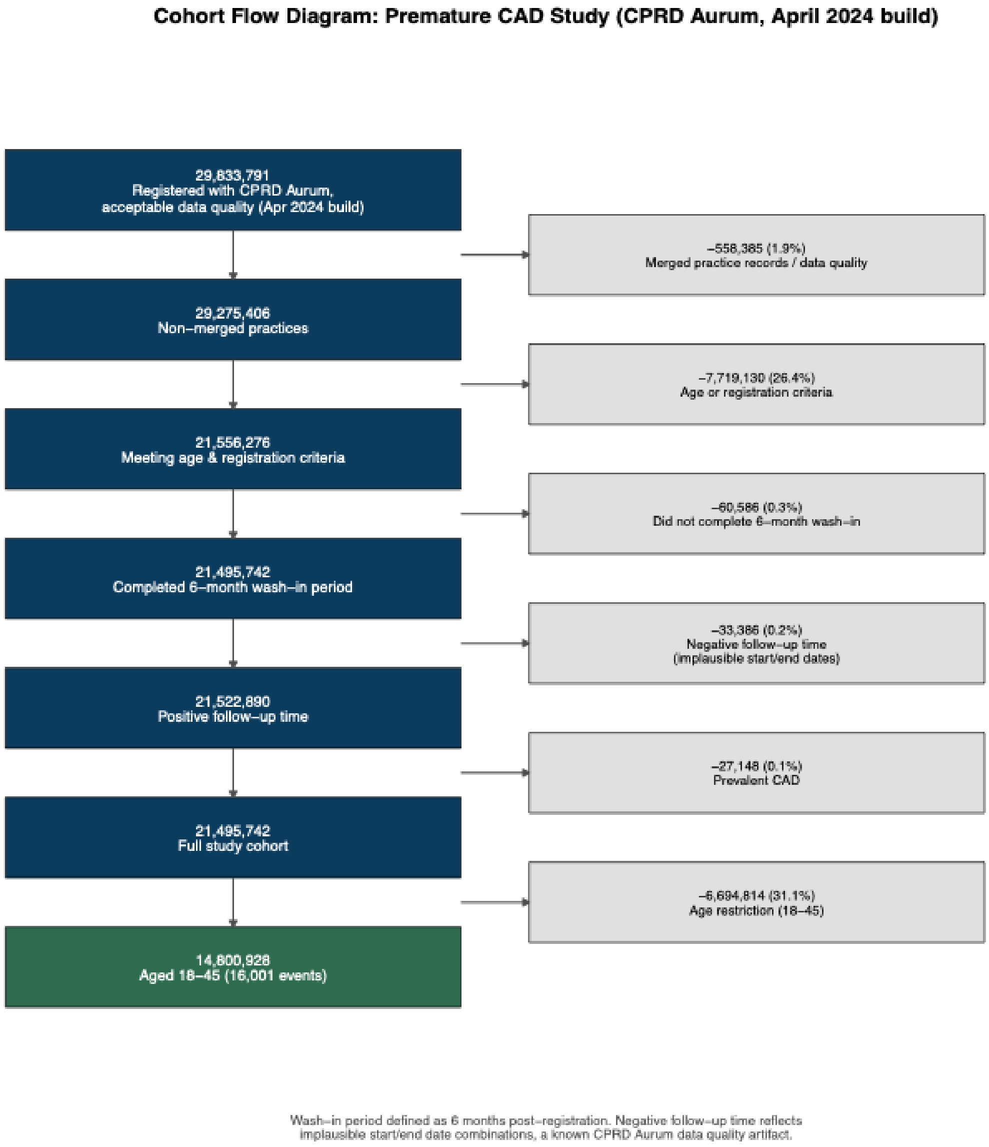

Figure S6: Flowchart for cohort creation.

| Data source | Content | Coverage period |
|---|---|---|
| CPRD Aurum (March 2024 build) | Primary care records, ~47 million patients, 1,596 English practices (EMIS) | 1995 – March 2024 |
| Hospital Episode Statistics (HES) Admitted Patient Care | Secondary care diagnoses and procedures | April 1997 – January 2022 |
| Index of Multiple Deprivation (IMD) | Patient-level socioeconomic deprivation | Linked at postcode level |
| ONS death registration | Date and cause of death | January 1998 – April 2021 |

Table S16: Data sources and linkage coverage

### Literature search strategy

Database: MEDLINE (via PubMed). Coverage: inception to 27 July 2026. Language: English only. Records retrieved: 1018.

| | |
|---|---|
| **Concept 1: Coronary disease** | |
| **#1** | "Coronary Artery Disease"[Mesh] OR "Myocardial Ischemia"[Mesh] OR "Myocardial Infarction"[Mesh] OR "Acute Coronary Syndrome"[Mesh] OR "Coronary Disease"[Mesh] |
| **#2** | (coronary artery disease[tiab] OR coronary heart disease[tiab] OR coronary disease[tiab] OR ischaemic heart disease[tiab] OR ischemic heart disease[tiab] OR myocardial infarction[tiab] OR myocardial ischaemia[tiab] OR myocardial ischemia[tiab] OR acute coronary syndrome[tiab] OR heart attack[tiab] OR coronary revascular*[tiab] OR percutaneous coronary intervention[tiab] OR coronary artery bypass[tiab] OR CAD[ti] OR CHD[ti] OR IHD[ti]) |
| **#3** | #1 OR #2 |
| **Concept 2: Premature / early-onset** | |
| **#4** | "Age of Onset"[Mesh] |
| **#5** | (premature[tiab] OR "early onset"[tiab] OR "early-onset"[tiab] OR "young adult*"[tiab]) |
| **#6** | #4 OR #5 |
| **Concept 3: Ethnicity / race / population groups** | |
| **#7** | "Ethnicity"[Mesh] OR "Racial Groups"[Mesh] OR "Asian People"[Mesh] OR "Black People"[Mesh] OR "Minority Groups"[Mesh] OR "Health Status Disparities"[Mesh] OR "Healthcare Disparities"[Mesh] OR "Emigrants and Immigrants"[Mesh] |
| **#8** | (ethnic*[tiab] OR South Asian*[tiab] OR Indian*[tiab] OR Pakistani*[tiab] OR Bangladeshi*[tiab] OR African*[tiab] OR Caribbean*[tiab] OR Afro-Caribbean*[tiab] OR "African Caribbean*"[tiab]) |
| **#9** | #7 OR #8 |
| **Combination** | |
| **#10** | #3 AND #6 AND #9 |
| **#11** | #10 AND English[Language] |
| **#12** | #11 NOT (Comment[pt] OR Editorial[pt] OR "Case Reports"[pt] OR Letter[pt]) |

*[Mesh] = Medical Subject Heading; [tiab] = title/abstract; [ti] = title only; [pt] = publication type; * = truncation.*
